# Preliminary Validation of a Structural Magnetic Resonance Imaging Metric for Tracking Dementia-Related Neurodegeneration and Future Decline

**DOI:** 10.1101/2022.11.10.22282162

**Authors:** Gavin T Kress, Emily S. Popa, Paul M Thompson, Susan Y Bookheimer, Sophia I Thomopoulos, Christopher RK Ching, Hong Zheng, David A. Merrill, Stella E Panos, Prabha Siddarth, Jennifer E Bramen

## Abstract

Current Alzheimer’s disease (AD) research has a major focus on validating and discovering noninvasive biomarkers that can detect AD, benchmark disease severity, and aid in testing the efficacy of interventions. Structural magnetic resonance imaging (sMRI) is a well-validated tool used in diagnosis and for monitoring disease progression in AD. Much of the sMRI literature centers around hippocampal and other medial temporal lobe structure atrophy, which are strongly associated with cognition and diagnosis. Because atrophy patterns are complex and vary by patient, researchers have made efforts to condense more brain information into validated metrics. Many of these methods use machine learning (ML), which can be difficult to interpret clinically, hampering clinical adoption. Here, we introduce a practical, clinically meaningful and interpretable index which we call an “AD-NeuroScore.” Our approach is automated and uses multiple regional brain volumes associated with cognitive decline. We used a modified Euclidean inspired distance function to calculate the differences between each participant and a cognitively normal (CN) older adult template, adjusting for intracranial volume, age, sex, and scanner model. Here we report validation results, including sensitivity to diagnosis (CN, mild cognitive impairment (MCI), and AD) and disease severity (Clinical Dementia Rating Scale Sum of Boxes (CDR-SB), Mini Mental State Exam (MMSE), and Alzheimer’s Disease Assessment Scale-Cognitive Subscale (ADAS-11) in 929 older adults (mean age=72.7 years, SD=6.3, Range=55.1-91.5, 50% Female) drawn from the Alzheimer’s Disease Neuroimaging Initiative (ADNI) study. To determine if AD-NeuroScore might be predictive of disease progression, we assessed the relationship between the calculated AD-NeuroScore at baseline and change in both diagnosis and disease severity scores at 12, 24, 36, and 48-months. We performed additional validation in all analyses, benchmarking AD-NeuroScore against adjusted hippocampal volume (AHV). We found that AD-NeuroScore was significantly associated with diagnosis and all disease severity scores at baseline. Associations between AD-NeuroScore and disease severity (CDR-SB and ADAS-11) were significantly stronger than with AHV. Baseline AD-NeuroScore was also associated with change in diagnosis and changes in disease severity scores at all time points. Performance was equivalent, or in some cases superior, to AHV. These early validation results suggest that AD-NeuroScore has the potential to be a clinically meaningful biomarker for dementia.

## 1. Introduction

The prevalence of Alzheimer’s disease (AD) is increasing with the aging population. According to the World Health Organization, there are an estimated 36 million patients with AD in 2021 with approximately 6.5 million new cases reported annually (WHO, 2021). AD is a progressive neurodegenerative disease characterized by cognitive decline and early atrophy within the medial temporal lobe (MTL) followed by progression to the rest of the brain. Neurodegeneration often precedes cognitive decline (Gómez-Isla et al., 1996a), making atrophy assessed with structural magnetic resonance imaging (sMRI) an ideal clinical biomarker for detecting AD early and forecasting future cognitive and functional deterioration. Detecting AD prior to the point of irreversible neurodegeneration could improve the efficacy of currently available treatments (Coupé et al., 2015a) and improvements to current quantitative sMRI would add value in both clinical and research settings.

At later stages of AD, additional and validated quantitative sMRI metrics could add clinical utility while monitoring disease progression. Neurocognitive assessments become more burdensome and less valuable as patients convert to moderate and severe AD. Regardless, there is still high clinical value in tracking a patient’s disease progression and anticipating future deterioration throughout all stages of illness. sMRI is a passive, relatively short and inexpensive measurement, making it an ideal modality for monitoring disease progression in general, but especially in later stage AD patients.

sMRI has wide adoption in both clinical and research settings. The majority of the sMRI literature centers around hippocampal or other MTL measures. The magnitude of hippocampal volume atrophy is strongly associated with cognition, diagnosis, and AD-etiology (Csernansky et al., 2004; Gosche et al., 2002; Jack et al., 2002; Silbert et al., 2003). Hippocampal shape (Achterberg et al., 2014), texture (Sørensen et al., 2016), grading (Coupé et al., 2015b) and subfield volumes (Khan et al., 2015) are all validated hippocampus-based markers of AD-related etiology. The MTL has generally been a focus due to its critical role in memory and because MTL atrophy is often detectable in cognitively normal (CN) individuals before symptoms of cognitive decline arise (Jack et al., 1997).

However, other parts of the brain also provide important information, especially in the differential diagnosis of mild cognitive impairment (MCI), AD, and frontotemporal lobe dementia (FTD), which have overlapping atrophy within the MTL (Rabinovici et al.,2007). A large body of research on AD-related etiology shows volume loss (Rabinovici et al., 2007) that quickly spreads from the MTL (Braak et al., 1997; Dickerson et al., 2001; Frisoni et al., 1999; Gómez-Isla et al., 1996b; Laakso et al., 1998, 1996; Rabinovici et al., 2007; Thompson et al., 2007, 2003; Vercelletto et al., 2002) to the parietal lobe (Jacobs et al., 2012a), where grey matter (GM) and white matter (WM) loss likely occurs relatively early in cortical progression and is linked functionally to important AD symptoms such as impaired memory retrieval, naming ability, attention, and executive function (Cabeza et al., 2008; Jacobs et al., 2012b; Lindeboom and Weinstein, 2004). Over time and with variability in the order and rate of decline amongst patients, neurodegeneration spreads to posterior temporal, lateral occipital (Rabinovici et al., 2007), and left frontal lobes as well as limbic structures including the thalamus, cingulate gyrus, and nucleus accumbens (Nie et al., 2017). Because the underlying pattern of atrophy is important to consider in each patient, clinical imaging reports that quantify regional atrophy are standard in many dementia clinics and regional volumes are common endpoints in clinical research.

Several medical imaging companies have implemented regional brain volume reports in clinical settings (Ahdidan et al., 2016; Brewer et al., 2009; Cavedo et al., 2022). These tools are helpful in understanding the overall pattern of neurodegeneration. However, the spatiotemporal pattern of atrophy varies widely in patients, and it is difficult to quickly assess a patient’s overall neurodegeneration from several numbers, reducing utility in diagnosis, which is most commonly done using neurocognitive assessment. Further, patients often receive scans at different facilities or on different scanners, and many of the current commercial tools do not account for these differences, affecting the validity of longitudinal follow-up reports. A valid, harmonized summary score with norms and cutoffs would be an important addition to existing, clinically-implemented regional brain volume reports. In research settings, several regions are frequently used as endpoints (e.g., hippocampal, entorhinal cortical, lateral ventricle, and whole brain volumes). Depending on the design and sample size of the study, reducing the number of regions to be considered as endpoints could enhance sensitivity. Additionally, a validated, harmonized score that could be compared across multiple research studies could be valuable for meta-analyses and help researchers build on each other’s findings.

Because atrophy patterns are complex and vary by patient, making the qualitative interpretation of several individual region of interest (ROI) results laborious, some researchers have condensed sMRI information into validated metrics for AD-related neurodegeneration. Early sMRI-based, multivariate analysis approaches have focused on the classification of CN and AD (Riciotti et al., 2012a, Vermuri, 2009). As the field has matured, researchers have aimed to classify more disease stages (i.e. CN, MCI, and AD) (Rallabandi et al., 2020; Popuri et al., 2020) as well as predict who will stabilize and who will decline (Coupé et al., 2015c; Ezzati et al., 2019; Popuri et al., 2020b). Most of these multivariate scoring methods use machine learning (Casanova et al., 2018; Diciotti et al., 2012a; Vermuri, 2009) and several combine sMRI with other imaging, biochemical, or clinical features to improve performance (Dukart et al., 2013; Salvatore et al., 2018; Vemuri et al., 2009a).

The objective of this study is to develop and validate a practical, clinically meaningful, and interpretable biomarker, which we call an “AD-NeuroScore” by summarizing AD-related neurodegeneration using only sMRI-based measurements. Our approach uses a vector of multiple sMRI ROI-based features that are associated with cognitive decline in an intuitive manner that is easy to implement. ROI-based features from only one type of image were selected because they are widely adopted by researchers and clinicians and because their significance is well understood. This is in contrast to multivariate models that successfully use machine learning approaches to generate probability scores (Casanova et al., 2018b; Diciotti et al., 2012b; Lu et al., 2021; Popuri et al., 2020b; V P.Subramanyam Rallabandi et al., 2020; Vemuri et al., 2009a) or more narrowly focused regional scores (Achterberg et al., 2014; Ahdidan et al., 2016; Brewer et al., 2009; Coupé et al., 2015d; Elahi et al., 2015; Sean M Nestor et al., 2008; Sørensen et al., 2016). Machine learning-based, black box models are powerful but have been have not yet been widely implemented in neurological practices. Model interpretability is an important barrier to clinical adoption despite the need for quality control in ROI-based approaches (Pinto et al., 2022). Further, while it is possible to understand the algorithms behind the ML models, this is not the same as having a clinically interpretable and meaningful metric. Here we developed an automated approach to assist in clinical practice, resulting in a summary score calculated using only well-understood sMRI regional volumes, where the relative contribution of each ROI is transparent. More narrowly focused ROI-based approaches are interpretable (Achterberg et al., 2014; Ahdidan et al., 2016; Archetti et al., 2021; Brewer et al., 2009; Casanova et al., 2013; Coupé et al., 2015d; Davatzikos et al., 2009; Sørensen et al., 2016), however, they, in some cases have difficulty capturing a significant portion of sMRI features due to the loss of information in the process of combining to a single metric. The biomarker we propose here, AD-NeuroScore, is automated, computationally inexpensive, and relies on widely adopted and validated sMRI features. We believe these attributes could enable widespread research and clinical deployment.

## 2. Methods

### 2.1. Participants

#### 2.1.1 Data Acquisition and Demographics

CN individuals and patients with MCI or AD diagnosis were drawn from the Alzheimer’s Disease Neuroimaging Initiative (ADNI) database (adni.loni.usc.edu) (Jack et al., 2008; Mueller et al., 2005a, 2005b). The ADNI is a global research study launched in 2003, primarily aimed at evaluating biological markers to measure the progression of MCI and early AD. Biomarkers will be derived from MRI, PET, and blood and cerebrospinal fluid (CSF)-based biospecimen assays and their discovery and validation are intended to facilitate the development of treatments to slow or halt AD progression. For up-to-date information, see www.adni-info.org.

All ADNI studies were conducted according to the Good Clinical Practice guidelines, the Declaration of Helsinki, the U.S. 45 Code of Federal Regulations (CFR) Part 46 and 21 CFR Part 50 – Protection of Human Subjects, and 21 CFR Part 56 - Institutional Review Boards. Written informed consent and HIPAA authorizations were obtained from all participants or authorized representatives prior to the conducting of protocol-specific procedures. The ADNI protocol was approved by the Institutional Review Boards of all participating institutions, listed in File 1 in Supplementary Materials (Mukherji et al., 2021).

In this work, a total of 1,619 subjects with available 3T, accelerated, T1-weighted MR images at baseline were collated from the ADNI-GO, ADNI-2, and ADNI-3 study phases; of these, 388 participants were discarded due to incomplete data used to calculate or validate the biomarker, including diagnosis (n=42), sex (n=45), age (n=8), scanner manufacturer (n=3), and at least one brain volume (n=298). Acquisition methods are detailed by Chow et al. (Chow et al., 2015). The remaining cohort of 1,231 individuals was comprised of 488 CN, 564 MCI, and 179 mild AD. Participants were diagnosed at baseline and reassessed at each study visit (Petersen et al., 2010). The study sample was subdivided into the following three cohorts: region of interest (ROI) selection, cognitively normal template creation, and experimental (see **Table 1** for a full description of all cohort demographics).

**Table 1.** Demographic Information. Full description of all cohort demographics at baseline. Total number of subjects (n), sex (by percentage female), age, and education in years are reported for each cohort in full (All), as well as for CN, MCI, and AD diagnostic subdivisions. The CN Template Cohort was comprised of only CN individuals. Values for age and education are summarized in the form mean (± standard deviation, min - max) and mean (± standard deviation), respectively.

| Dataset | Clinical Measure | Group |  |  |  |
| --- | --- | --- | --- | --- | --- |
|  |  | CN | MCI | AD | All |
| ROI Selection | n | 50 | 50 | 50 | 150 |
|  | Subjects (% Female) | 62 | 48 | 42 | 51 |
| | Age | 68.6 (±6.5, 55.1-88.7) | 71.9 (±8.6, 55.2-88.7) | 74.9 (±7.3, 55.3-89.7) | 71.8 (±7.9, 55.1-89.9) |
| | Education | 17.2 (±1.9) | 16.2 (±2.3) | 15.7 (±2.4) | 16.4 (±2.3) |
| Template | n | 152 |  |  |  |
|  | Subjects (% Female) | 50 |  |  |  |
| | Age | 71.7 (±5.9, 55.6-85.3) | | | |
| | Education | 16.9 (±2.2) | | | |
| Experimental | n | 286 | 514 | 129 | 929 |
|  | Subjects (% Female) | 64 | 45 | 43 | 50 |
| | Age | 72.5 (±6.3, 56.3-90.2) | 72.1 (±7.5, 55.1-91.5) | 74.7 (±8.3, 55.7-90.4) | 72.6 (±7.3, 55.1-91.5) |
| | Education | 16.5 (±2.5) | 16.2 (±2.6) | 15.7 (±2.6) | 16.2 (±2.6) |

#### 2.1.2 Region of Interest Selection Cohort

To determine the regional volumes that were associated with cognitive impairment, a sample of 150 age- and sex-matched individuals from the overall participant pool were pseudo-randomly selected using the numpy random module (Harris et al., 2020) with equivalent proportions of each diagnostic category (see **Table 1** for a full description of ROI selection cohort demographics) and grouped into a cohort in order to quantitatively determine ROI inclusion and weighting (see **Methods 2.3.2. ROI Selection)**.

#### 2.1.3 Cognitively Normal Template Cohort

A sample of cognitively normal, age- and sex-matched individuals were randomly selected from the remaining participant pool to generate a CN template. The purpose of this template was to represent the average, healthy older adult brain (see **Table 1** for a full description of CN template demographics). The sample size for this cohort was set to 152 individuals, to match that which was drawn from a normative adult population to construct the MNI152 template, a classic and widely adopted structural brain atlas based on the group-wise registration of 152 3D, T1-weighted MR images (Mazziotta et al., 1995a, 1995b).

#### 2.1.4 Experimental Cohort

The remaining 929 participants were grouped into an experimental cohort for sMRI metric extraction and testing (see **Table 1** for a full description of experimental cohort demographics). Baseline analyses were conducted for all 929 participants of the experimental cohort.

Longitudinal analyses were conducted on all participants who had follow-up data available. These participants were classified as those who worsened in diagnosis (denoted as Diagnosis_decline_) and those who did not change in diagnosis or, in rare cases, improved (denoted as Diagnosis_stable_), based on their diagnosis at 12, 24, 36 and 48-month follow-up sessions. Diagnosis_decline_ is thus composed of those who declined from CN to MCI or CN to AD (CN_decline_) and from MCI to AD (MCI_decline_); Diagnosis_stable_ is the group who either did not change in diagnosis or improved (composed of MCI_stable_, CN_stable_, and AD_stable_). There was no later stage diagnostic category than AD in this sample, and therefore all AD participants were AD_stable_, with no AD_decline_ group (see **Figure 1** for a flowchart detailing the construction of longitudinal cohorts).

**Figure 1.**
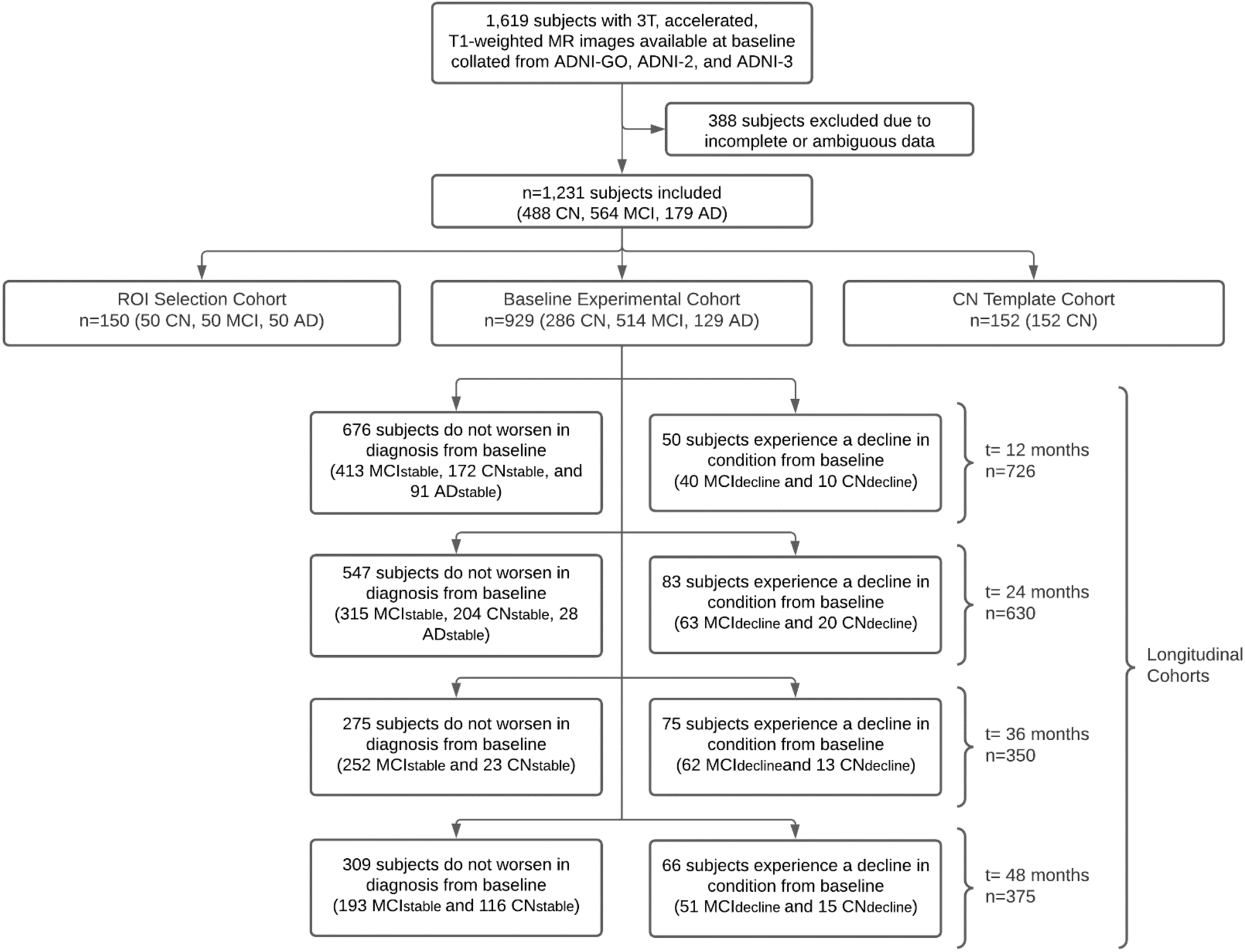
Flowchart visualizing baseline allocation of subjects into the 3 experimental cohorts (ROI Selection, CN Template, and Experimental) and longitudinal construction of the stable and decline groups at 12, 24, 36, and 48-months. Patients whose diagnosis progressed relative to baseline were classified into a Diagnosis (decline) group, while those who did not worsen were categorized as Diagnosis (stable) for each respective time point.

### 2.2 Neurocognitive Assessments

Participant scores on the Clinical Dementia Rating Scale Sum of Boxes (CDR-SB), Alzheimer’s Disease Assessment Scale Cognitive Subscale (ADAS-11), and Mini-Mental State Exam (MMSE) were collated for all experimental cohort participants with scores available at each time point. The original ADAS-Cog (Rosen et al., 1984), referred to as the ADAS-11, is comprised of 11 subtests whose scores are summed (unweighted) to generate a total raw score from 0 to 70. Higher scores on the ADAS-11 correspond with a larger number of errors and poorer performance, as rated by clinicians (Grochowalski et al., 2015). The MMSE is a short, 11 items questionnaire routinely used to measure cognitive impairment (Folstein et al., 1975). The MMSE has six subdomains: time orientation, place orientation, registration, attention, recall, language, and visual construction and yields a score from 0 to 30, with lower scores representing greater cognitive dysfunction The CDR-SB (Balsis et al., 2015; Hughes et al., 1982; Morris, 1993; O’Bryant et al., 2008) similarly measures cognitive function, along with functional ability, as rated by clinicians across six domains (also referred to as boxes): memory, orientation, judgement and problem-solving, community affairs, home and hobbies, and personal care. Individual scores for each domain (or box) are summed to produce a total score ranging from 0 to 18, where higher scores stage more severe dementia (O’Bryant et al., 2008)

### 2.3 AD-NeuroScore

#### 2.3.1

Cortical reconstruction and volumetric segmentation of the 3T, accelerated, T1-weighted MR images was performed with version 7.1 of the Freesurfer image analysis suite, which is documented and freely available for download online (http://surfer.nmr.mgh.harvard.edu/). The details of these procedures are described in previous publications (Dale et al., 1999; Desikan et al., 2006; Fischl et al., 2004a, 2004b, 2002, 2001, 1999a, 1999b; Jovicich et al., 2006; Reuter et al., 2010; Segonne et al., 2007, 2004). With these methods, eighty-four cortical and subcortical regional volumes were estimated at baseline for each subject. Additionally, results were reviewed using the ENIGMA structural imaging quality control protocols (http://enigma.usc.edu/) (Stein et al., 2012).

#### 2.3.2. ROI Selection

In order to determine the brain regions most sensitive to diagnosis at baseline, we first performed an Analysis of Variance (ANOVA) in our ROI selection cohort to test for an effect of diagnosis for each of the 84 regions (using a Bonferroni-corrected alpha of 0.05/84; see **Supplementary File 2** for a complete list of the 84 cortical and subcortical regions tested). Forty-one regions were found to be significant. All these regions were consistent with the existing literature (Harper et al., 2017; Yin et al., 2013; Zanchi et al., 2017) and were used to compute AD-NeuroScore (**Table 2**).

**Table 2.** Significant ROIs by Z-Score Ranking and CN Template Values. Resulting 41 significant regions of interest (ROIs) extracted by performing ANOVA for each of the 84 regions in the ROI selection cohort are reported in the above table along with corresponding z scores. Significance was established based on an alpha=0.05, Bonferroni corrected. Structures are identified by both the FreeSurfer version 7.1 ROI labels and conventional anatomical names. Mean volumes and standard deviations (SD) of the CN template cohort are included for each respective region.

| FreeSurfer ROI Label | Anatomical Name | Z | CN Template Mean Vol (±SD) (mm<sup>3</sup>) |
| --- | --- | --- | --- |
| Lhippo | Left Hippocampus | 9.12 | 0.48 (±0.74) |
| Lamyg | Left Amygdala | 8.43 | 0.60 (±0.79) |
| Ramyg | Right Amygdala | 8.37 | 0.54 (±0.80) |
| Rhippo | Right Hippocampus | 7.82 | 0.45 (±0.71) |
| L_middletemporal_grayvol | Left Middle Temporal Lobe | 6.53 | 0.40 (±0.88) |
| L_fusiform_grayvol | Left Fusiform Gyrus | 6.20 | 0.27 (±0.82) |
| L_inferiorparietal_grayvol | Left Inferior Parietal Lobe | 6.11 | 0.33 (±0.90) |
| R_middletemporal_grayvol | Right Middle Temporal Lobe | 6.12 | 0.42 (±0.92) |
| L_inferiortemporal_grayvol | Left Inferior Temporal Lobe | 5.88 | 0.36 (±0.85) |
| L_precuneus_grayvol | Left Precuneus | 5.81 | 0.26 (±0.91) |
| L_rostralmiddlefrontal_grayvol | Left Rostral Middle Frontal Lobe | 5.69 | 0.16 (±1.06) |
| R_inferiortemporal_grayvol | Right Inferior Temporal Lobe | 5.54 | 0.27 (±0.87) |
| R_precuneus_grayvol | Right Precuneus | 5.50 | 0.34 (±0.93) |
| R_inferiorparietal_grayvol | Right Inferior Parietal Lobe | 5.48 | 0.33 (±0.88) |
| R_superiortemporal_grayvol | Right Superior Temporal Lobe | 5.44 | 0.35 (±0.94) |
| R_entorhinal_grayvol | Right Entorhinal Cortex | 5.39 | 0.12 (±0.77) |
| Lthal | Left Thalamus | 5.17 | 0.33 (±1.01) |
| R_lateralorbitofrontal_grayvol | Right Lateral Orbitofrontal Cortex | 5.02 | 0.14 (±0.91) |
| L_superiortemporal_grayvol | Left Superior Temporal Lobe | 5.01 | 0.28 (±0.87) |
| L_insula_grayvol | Left Insula | 4.86 | 0.19 (±0.86) |
| L_bankssts_grayvol | Left Banks of the Superior Temporal Sulcus | 4.75 | 0.12 (±0.93) |
| L_lateralorbitofrontal_grayvol | Left Lateral Orbitofrontal Cortex | 4.67 | 0.26 (±0.89) |
| L_isthmuscingulate_grayvol | Left Isthmus of the Cingulate Gyrus | 4.66 | 0.20 (±1.04) |
| R_parahippocampal_grayvol | Right Parahippocampal Gyrus | 4.64 | 0.21 (±0.91) |
| Raccumb | Right Nucleus Accumbens | 4.58 | 0.33 (±0.88) |
| L_entorhinal_grayvol | Left Entorhinal Cortex | 4.56 | 0.23 (±0.74) |
| R_rostralmiddlefrontal_grayvol | Right Rostral Middle Frontal Gyrus | 4.50 | 0.18 (±1.04) |
| L_superiorparietal_grayvol | Left Superior Parietal Lobe | 4.44 | 0.14 (±0.87) |
| Rthal | Right Thalamus | 4.42 | 0.31 (±1.02) |
| L_superiorfrontal_grayvol | Left Superior Frontal Gyrus | 4.38 | 0.17 (±0.89) |
| R_fusiform_grayvol | Right Fusiform Gyrus | 4.25 | 0.23 (±0.88) |
| R_insula_grayvol | Right Insula | 4.14 | 0.21 (±0.93) |
| L_lateraloccipital_grayvol | Left Lateral Occipital Cortex | 4.08 | 0.31 (±0.87) |
| R_superiorfrontal_grayvol | Right Superior Frontal Gyrus | 3.96 | 0.16 (±0.99) |
| L_posteriorcingulate_grayvol | Left Posterior Cingulate Cortex | 3.86 | 0.29 (±0.98) |
| R_isthmuscingulate_grayvol | Right Isthmus of the Cingulate Gyrus | 3.78 | 0.13 (±0.87) |
| R_superiorparietal_grayvol | Right Superior Parietal Lobe | 3.75 | 0.26 (±1.00) |
| R_parsorbitalis_grayvol | Pars Orbitalis | 3.70 | 0.19 (±0.92) |
| R_posteriorcingulate_grayvol | Right Posterior Cingulate Cortex | 3.60 | 0.24 (±0.90) |
| Laccumb | Left Nucleus Accumbens | 3.60 | 0.45 (±1.04) |
| R_lateraloccipital_grayvol | Right Lateral Occipital Cortex | 3.47 | 0.24 (±0.83) |

#### 2.4.1. Cognitively Normal Template

To generate a CN template vector representative of the healthy average older adult brain, each volume that was determined from the ROI selection process was averaged across all CN template cohort participants (see **Table 2**). Data harmonization procedures were applied to each region in the CN template vector (see **Methods 2.4.2. Data Harmonization).** This CN template vector had the same dimensions as vectors extracted from experimental cohort participants. These two vectors were used to compute the distance metrics evaluated in this work (see Methods 2.4.3. Calculating the Z-Weighted Euclidean Distance).

#### 2.4.2. Data Harmonization

In order to account for factors such as interindividual variations in head size, age, sex, and MRI acquisition features like scanner model and manufacturer, we utilized a w-score, a data harmonization approach previously validated on sMRI data (Ma et al., 2019; Popuri et al., 2020a). This approach uses a generalized linear model (GLM) framework, where the structural volume for a given region of each participant is modeled as the linear combination of all covariates as shown in Equation (1):

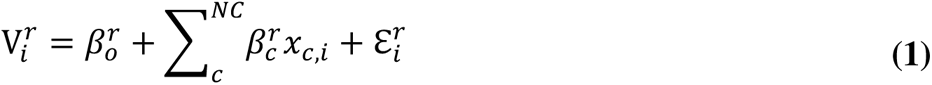

Here, the volume 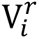 of region r for subject i, is modeled as the sum over NC covariates (age, sex, scanner model, and total ICV) of coefficient 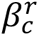, for covariate c, multiplied by the value of the covariate, 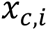, plus the residual term, 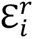.

To compute the w-score, 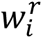, also known as the standardized residual, we took the z-transformation of the residual term from Eq. (1), 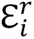, as shown in Equation (2):

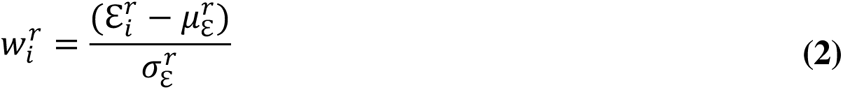

where 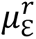 and 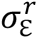 represent the mean and standard deviation, respectively, of the residual term for a given subject’s regional brain volume. In this way, regional volumes used to calculate AD-NeuroScore were adjusted for the effects of age, sex, head size, and scanner model.

#### 2.4.3 Calculating the Z-Weighted Euclidean Distance

To compute the difference between each experimental cohort participant and the CN template, we employed a novel modified Euclidean inspired distance function which we call the Z-weighted Euclidean distance (ZWE). A traditional Euclidean distance metric is calculated by treating the list of significant, harmonized regional volumes as a vector in n-dimensional space, where n is the number of regions, and computing the Euclidean distance between each participant and the CN template vectors. Our ZWE distance function differed only in that each region was multiplied by a weight resulting from each region’s level of significance (z-score) as determined during ROI selection process as shown in Equation (3):

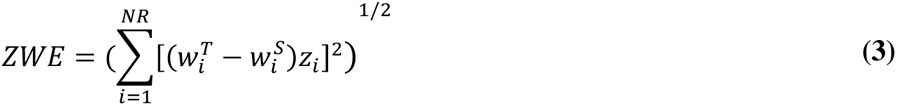

where *Z_i_* refers to the average of the z-scores associated with the p-values across the three possible pairwise comparisons (CN vs MCI, CN vs AD, and MCI vs AD) for a given region, i, and NR denotes the total number of regions. The ZWE distance was computed between the harmonized residual of each subject, 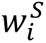, and the CN template, 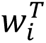. Thus, the more significant a region was in ROI-selection, the greater its contribution to the computed AD-NeuroScore. The concept of weighing multivariate sums by measures of significance has been used in a number of applications since its inception in the proposal of Hotelling’s T^2^ distance in 1947 (Gutman et al., 2013; Hotelling, 1947; Hua et al., 2013). The significant ROIs listed in **Table 2** are visualized in **Figure 2**, with a color scale indicating respective *Z_i_* weighting.

**Figure 2.**
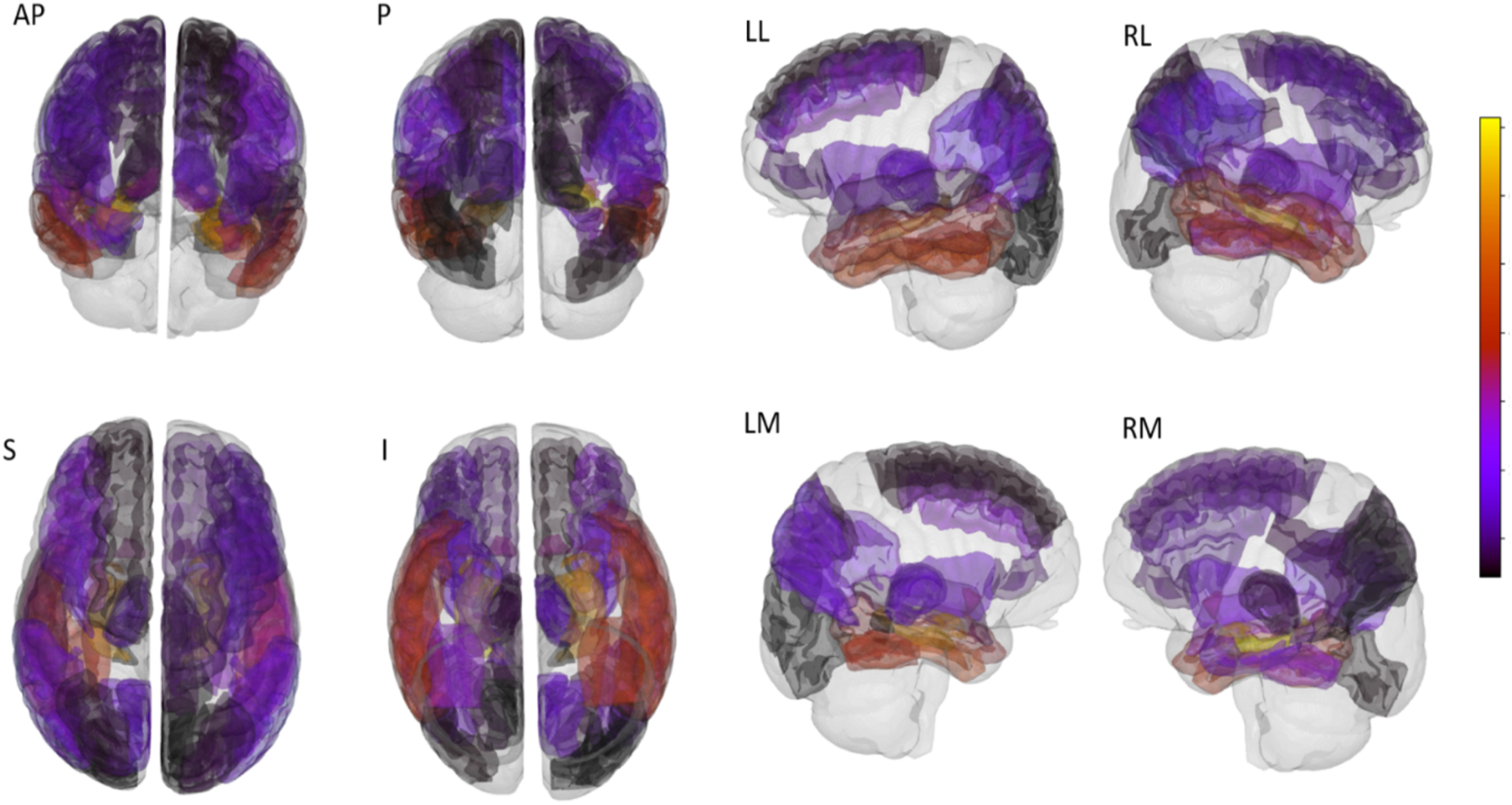
The 41 significant regions of interest (ROIs) extracted by performing ANOVA for each of the 84 regions in the ROI selection cohort are visualized above on the Allen 500-micron Human Brain Atlas. The color scale depicts the z-score-based weighting of each ROI in AD-NeuroScore (ADNS), as described in Section 2.4.4. The Brainrender python library (https://github.com/brainglobe/brainrender) was used to create this figure. The following letters denote anatomical orientation: Anterior (A), Posterior (P), Superior (S), Inferior (I), Left Lateral Surface (LL), Left Medial Surface (LM), Right Lateral Surface (RL), Right Medial Surface (RM).

We investigated several other methods for computing the difference between each experimental cohort subject and the CN template. Each of these algorithms similarly involved computing a distance function between like objects constructed from the harmonized, significant regional brain volumes, which we determined in the ROI selection step. Generally, the distance functions fell into one of two categories (curves and points). For the distance functions that computed distances between curves, in addition to projecting the curve in one dimensional space, the k-false nearest neighbors method was implemented to embed each curve in n-dimensions (see **Supplementary File 3** for further details). The functions which computed a distance between curves included the Fréchet distance and the Hausdorff distance. The Fréchet Distance is one method used to quantify the similarity between two curves or sets of points in space, with an emphasis on the location and ordering of points (Dumitrescu and Rote, 2004). Similarly, the Hausdorff distance measures how far two subspaces of a metric space are from each other; however, it does not account for the flow or order of points (Maiseli, 2021). The vector-based distance functions included the Euclidean and ZWE distance. Further details along with equations modeling these mathematical distances are included in **Supplemental File S3**.

Statistical analyses tested the performance of each distance metric based on sensitivity to diagnosis, disease severity, and progression (See **Tables 1-3** in **Supplementary Materials**). All metrics evaluated were benchmarked using AHV, an NIA-AA diagnostic biomarker for Alzheimer’s disease (Jack et al., 2018) (**Supplemental Table 1**). The optimal biomarker, the ZWE distance, was selected as our “AD-NeuroScore.” (See **2.5 Validation Procedures**).

**Table 3.** Baseline Results for AD-NeuroScore Sensitivity to Diagnosis. Sensitivity to diagnosis assessed using pairwise, two-tailed t-tests performed for each possible diagnostic group comparison. Resulting z-scores, effect sizes (Cohen’s *d*) with 95% confidence intervals (CI), and AUC-ROC values with 95% CI are included. Results using AHV are included for benchmarking. Significant results from group comparisons are denoted by * to indicate p<0.05, ** to indicate p<0.01, and *** to indicate p<0.001, Holm-Bonferroni corrected.

| Group Comparison | Metric | AUC [95% CI] | Cohen's *d* [95% CI] | z-score |
| --- | --- | --- | --- | --- |
| CN vs MCI | AD-NeuroScore | 0.66 [0.59, 0.73] | -0.56 [-0.59, -0.53]*** | -7.46 |
|  | AHV | 0.65 [0.58, 0.72] | 0.55 [0.52, 0.58]*** | -7.27 |
| CN vs AD | AD-NeuroScore | 0.91 [0.86, 0.96] | -2.06 [-2.24, -1.88]*** | -16.37 |
|  | AHV | 0.88 [0.81, 0.95] | 1.72 [1.55, 1.89]*** | -14.27 |
| MCI vs AD | AD-NeuroScore | 0.80 [0.71, 0.89] | -1.16 [-1.19, -1.14]*** | -11.23 |
|  | AHV | 0.76 [0.67, 0.84] | 0.92 [0.90, 0.94]*** | -9.04 |

### 2.5. Validation Procedures

#### 2.5.1. Baseline Validation Procedures

At baseline, we evaluated sensitivity to diagnosis of AD-NeuroScore by calculating the area under the Receiver Operating Characteristic curve (AUC-ROC) and the associated 95% confidence intervals, using a logistic regression model for each pairwise comparison of diagnostic groups (CN vs MCI, MCI vs AD, and CN vs AD). Sensitivity to diagnosis was further assessed using pairwise, two-tailed t-tests for each possible diagnostic group comparison, using a Holm-Bonferroni-corrected alpha of 0.05. Results were then converted to z-scores and Cohen’s *d* was calculated to indicate effect size.

Baseline associations of AD-NeuroScore and AHV with disease severity, operationalized as MMSE, ADAS-11, and CDR-SB scores, were tested using linear regression, both in the overall baseline experimental cohort and in each diagnostic sub-group. In addition to examining the significance of the slope using a Holm-Bonferroni-corrected alpha of 0.05, we also estimated the correlation coefficients and compared the performance of AD-NeuroScore and AHV by conducting a Fisher’s z-test of the z-transformed correlation coefficients, using the Holm-Bonferroni method to adjust for the 3 comparisons at baseline.

#### 2.5.2. Longitudinal Validation Procedures

To determine if AD-NeuroScore might be predictive of disease progression, we assessed the relationship between AD-NeuroScore at baseline and both the change in diagnosis and change in disease severity at 12, 24, 36, and 48 months. Logistic regression was used to examine whether baseline AD-NeuroScore was predictive of change in diagnosis (Diagnosis_decline_ vs. Diagnosis_stable_); AUC-ROC (and associated 95% CI) was used as a metric of the predictive ability. We also compared the baseline distribution of AD-NeuroScore between Diagnosis_decline_ and Diagnosis_stable_ groups using pairwise, two-tailed t-tests in the full experimental cohort and subsequently further stratified by baseline diagnosis (MCI or CN) to investigate if the ability to predict decline is driven by a specific patient population. Similar to the baseline validation procedures, the results for all comparisons were converted to z-scores, and Cohen’s *d* was calculated to assess effect size.

Longitudinal association with disease severity was tested using linear regression between baseline metric scores and the change in the neuropsychological assessment scores (MMSE, ADAS-11, and CDR-SB) from baseline, at each respective longitudinal session. Significance was assessed using a Holm-Bonferroni-corrected alpha of 0.05. All absent longitudinal comparisons were excluded due to insufficient sample size.

### 2.6. Validation Using Clinically Implemented ROI

To validate AD-NeuroScore in the context of the alternative imaging analysis tools frequently used by and accessible to clinicians, a vector of Neuroreader®-analagous ROI volumes was approximated by transforming the FreeSurfer results into a less granular vector of regions closely matching the Neuroreader® atlas. To investigate how these changes might impact biomarker performance, 80 of the 84 brain regions estimated by FreeSurfer were surjectively mapped to construct the 22 Neuroreader® ROI structures. Four of the eighty-four FreeSurfer-estimated regions, the left and right nucleus accumbens and insula, had no corresponding Neuroreader® ROI, and were thus dropped from this branch of the analysis. The calculation and validation methods used to compute and validate the Freesurfer-based AD-NeuroScore were applied to the pseudo-Neuroreader® ROI vector (**Supplementary Tables S4-S7**).

### 2.7. Data Availability

Full, open access to all de-identified ADNI imaging and clinical data is publicly and freely available to individuals who register with the ADNI and agree to the conditions in the “ADNI Data Use Agreement,” upon approval of a request that includes the proposed analysis and the named lead investigator (contact via http://adni.loni.usc.edu/data-samples/access-data/). Additional details about the ADNI data acquisition and sharing policies can be found at http://adni.loni.usc.edu/wp-content/uploads/how_to_apply/ADNI_DSP_Policy.pdf.

The code supporting the findings of this study is openly available in [repository name: “AD-NeuroScore”] at https://github.com/jbramen/AD-NeuroScore. The Python programming language (version 3.9) was used for all analyses (Python Software Foundation, https://www.python.org/).

## 3. Results

### 3.1. Baseline Validation

#### 3.1.1. Baseline Sensitivity to Diagnosis

AD-NeuroScore was significantly associated with diagnosis at baseline (p<0.001 for all comparisons, Holm-Bonferroni corrected; **Table 3**). AD-NeuroScore performed best at distinguishing AD from other groups (CN and MCI) and least well at distinguishing CN and MCI participants. AD-NeuroScore performed as well as our benchmark, AHV (p<0.001 for all comparisons, Holm-Bonferroni corrected), in this cross-sectional validation (**Table 3**). AUC-ROC values of AD-NeuroScore and AHV were similar across all group comparisons. Visual inspection of the overlaid AD-NeuroScore and AHV AUC-ROC curves for AD comparisons (CN vs AD and MCI vs AD) indicated that at low false positive rates, the AD-NeuroScore true positive rate tended to be higher (**Figure 3**). Examining the distribution of AD-NeuroScore and AHV for each diagnostic group at baseline (**Figure 4**), AD-NeuroScore qualitatively demonstrated greater separation between diagnostic category medians, with more centrally concentrated distributions and longer tails in CN and MCI groups at the edges of the distribution.

**Figure 3.**
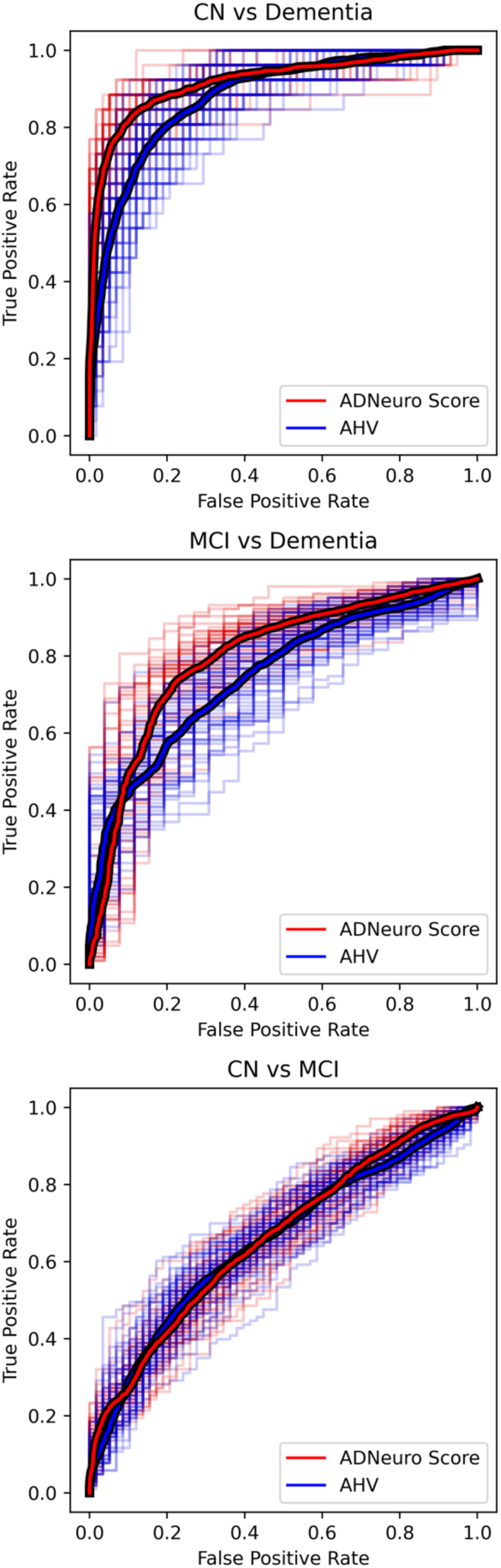
Overlaid AUC-ROC curves visualizing baseline classification performance of AD-NeuroScore and AHV across the 3 diagnostic group comparisons.

**Figure 4.**
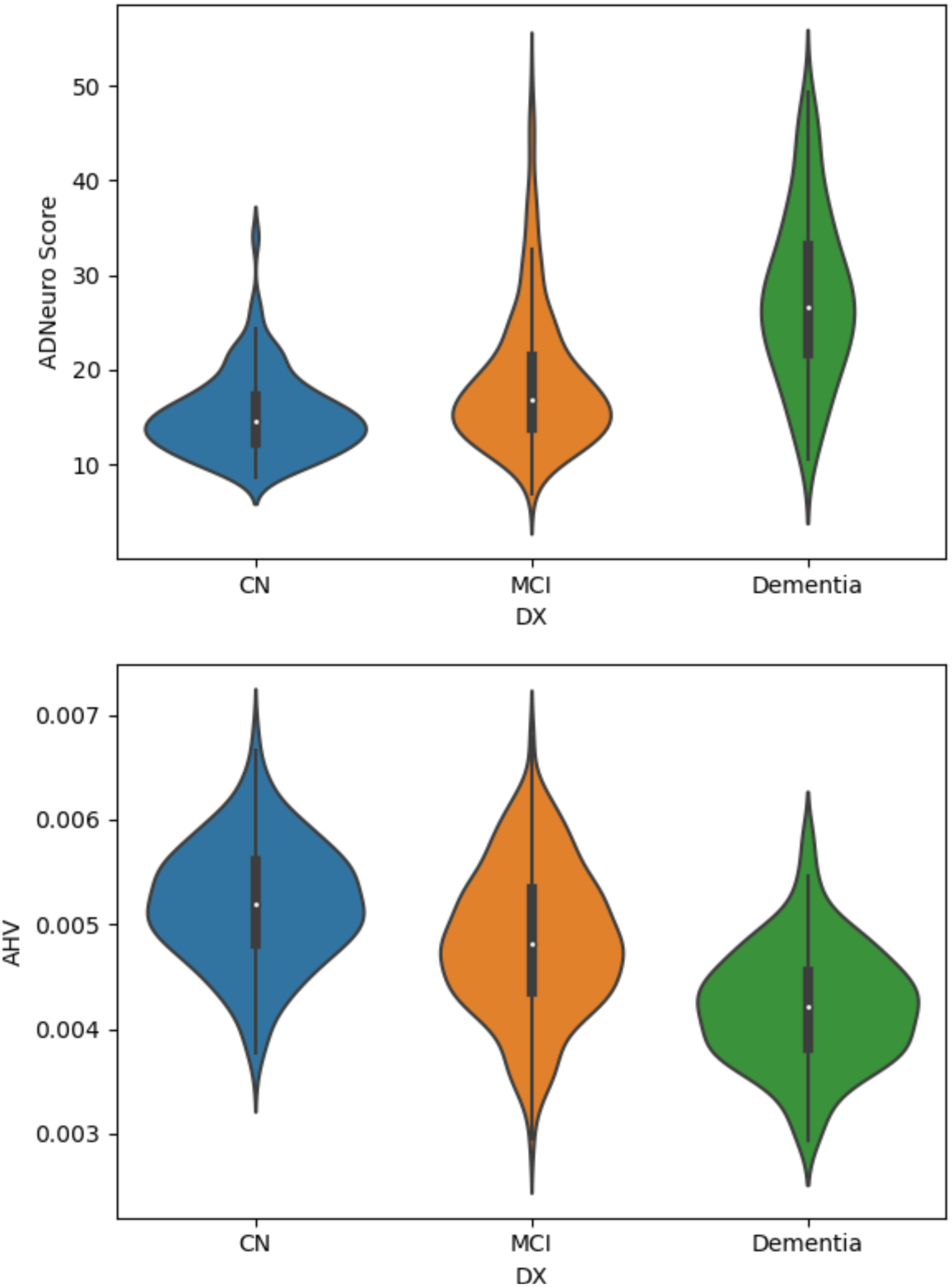
Violin plots depicting the distribution of AD-NeuroScore and AHV for each diagnostic group at baseline.

#### 3.1.2. Baseline Association with Disease Severity (MMSE, ADAS-11, and CDR-SB)

In the overall baseline experimental cohort, we found that AD-NeuroScore was significantly associated with disease severity, as measured by MMSE, ADAS-11, and CDR-SB scores (**Table 4**; **Figure 5**; all p-values < 0.001, Holm-Bonferroni corrected). In sub-analyses stratified by baseline diagnosis, we found significant associations between AD-NeuroScore and metrics of disease severity in participants with MCI (MMSE, ADAS-11, and CDR-SB) and AD (ADAS-11, and CDR-SB). Conversely, we found no significant association with disease severity in CN individuals.

**Figure 5.**
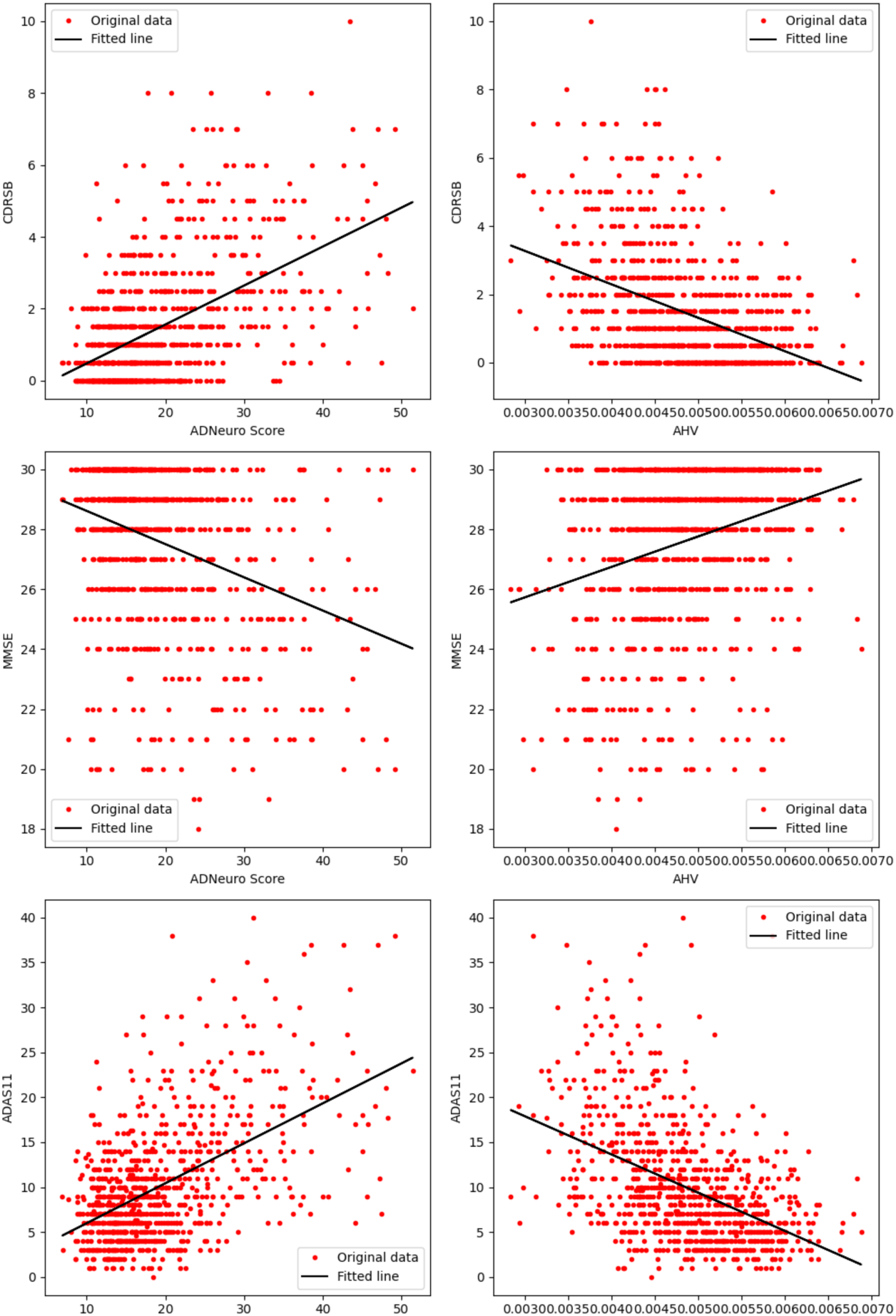
Correlations between each metric – AD-NeuroScore and AHV, with CDR-SB, MMSE, and ADAS-11 at baseline.

**Table 4.** Relationship Between AD-NeuroScore and Disease Severity. Baseline rows include results from cross-sectional analysis of AD-NeuroScore and disease severity, operationalized as MMSE, ADAS-11, and CDR-SB scores. All other rows include results from longitudinal analysis of baseline AD-NeuroScore and change in disease severity scores at 12, 24, 36, and 48 months. Testing was performed with linear regression in the overall experimental cohort and in each diagnostic sub-group. Performance of AHV is included for benchmarking. Significant associations are indicated by * for p<0.05, ** for p<0.01, and *** for p<0.001, Holm-Bonferroni corrected.

| Class | Time | Metric | I |  |  | II |  |  |
| --- | --- | --- | --- | --- | --- | --- | --- | --- |
|  |  |  | CDR-SB | MMSE | ADAS-11 | CDR-SB | MMSE | ADAS-11 |
| All | Baseline | AD-NeuroScore | 0.52*** | -0.34*** | 0.53*** | 926 | 926 | 926 |
|  |  | AHV | -0.43*** | 0.28*** | -0.47*** | 926 | 926 | 926 |
|  | 12 | AD-NeuroScore | -0.27*** | 0.21*** | -0.23*** | 525 | 697 | 721 |
|  |  | AHV | 0.23*** | -0.16*** | 0.19*** | 525 | 697 | 721 |
|  | 24 | AD-NeuroScore | -0.40*** | 0.27*** | -0.39*** | 402 | 614 | 626 |
|  |  | AHV | 0.32*** | -0.19*** | 0.26*** | 402 | 614 | 626 |
|  | 36 | AD-NeuroScore | -0.38*** | 0.21*** | -0.31*** | 285 | 332 | 345 |
|  |  | AHV | 0.34*** | -0.17** | 0.26*** | 285 | 332 | 345 |
|  | 48 | AD-NeuroScore | -0.42*** | 0.26*** | -0.31*** | 205 | 369 | 375 |
|  |  | AHV | 0.35*** | -0.19*** | 0.26*** | 205 | 369 | 375 |
| CN | Baseline | AD-NeuroScore | -0.08 | -0.01 | 0.10 | 286 | 286 | 286 |
|  |  | AHV | 0.00 | -0.01 | -0.17 | 286 | 286 | 286 |
|  | 12 | AD-NeuroScore | -0.81* | -0.02 | 0.09 | 13 | 183 | 180 |
|  |  | AHV | 0.53 | -0.03 | 0.01 | 13 | 183 | 180 |
|  | 24 | AD-NeuroScore | -0.69* | 0.02 | -0.09 | 17 | 220 | 221 |
|  |  | AHV | 0.45 | -0.08 | 0.05 | 17 | 220 | 221 |
|  | 36 | AD-NeuroScore | - | -0.06 | 0.01 | 1 | 44 | 36 |
|  |  | AHV | - | -0.19 | 0.03 | 1 | 44 | 36 |
|  | 48 | AD-NeuroScore | -0.12 | 0.00 | 0.01 | 5 | 126 | 133 |
|  |  | AHV | 0.52 | -0.21 | -0.01 | 5 | 126 | 133 |
| MCI | Baseline | AD-NeuroScore | 0.23*** | -0.09* | 0.33*** | 511 | 511 | 511 |
|  |  | AHV | -0.22*** | 0.11* | -0.35*** | 511 | 511 | 511 |
|  | 12 | AD-NeuroScore | -0.30*** | 0.21*** | -0.19*** | 418 | 423 | 452 |
|  |  | AHV | 0.23*** | -0.13* | 0.17** | 418 | 423 | 452 |
|  | 24 | AD-NeuroScore | -0.39*** | 0.28*** | -0.34*** | 357 | 357 | 377 |
|  |  | AHV | 0.32*** | -0.15* | 0.25*** | 357 | 357 | 377 |
|  | 36 | AD-NeuroScore | -0.38*** | 0.24*** | -0.31*** | 284 | 273 | 309 |
|  |  | AHV | 0.35*** | -0.16* | 0.27*** | 284 | 273 | 309 |
|  | 48 | AD-NeuroScore | -0.42*** | 0.30*** | -0.31*** | 199 | 226 | 242 |
|  |  | AHV | 0.35*** | -0.19* | 0.28*** | 199 | 226 | 242 |
| AD | Baseline | AD-NeuroScore | 0.27* | -0.17 | 0.33** | 129 | 129 | 129 |
|  |  | AHV | -0.08 | 0.08 | -0.14 | 129 | 129 | 129 |
|  | 12 | AD-NeuroScore | 0.09 | 0.04 | 0.00 | 94 | 91 | 89 |
|  |  | AHV | -0.08 | -0.03 | -0.22 | 94 | 91 | 89 |
|  | 24 | AD-NeuroScore | 0.03 | 0.16 | -0.36 | 28 | 37 | 28 |
|  |  | AHV | -0.34 | -0.10 | -0.20 | 28 | 37 | 28 |
|  | 36 | AD-NeuroScore | - | -0.13 | - | 0 | 15 | 0 |
|  |  | AHV | - | -0.09 | - | 0 | 15 | 0 |
|  | 48 | AD-NeuroScore | - | 0.28 | - | 1 | 17 | 0 |
|  |  | AHV | - | -0.11 | - | 1 | 17 | 0 |

AD-NeuroScore generally performed as well or better than our benchmark, AHV, in this cross-sectional validation using the overall baseline experimental cohort (**Table 4**; **Figure 5**). Results from z-tests conducted using Fisher z-transformed correlation coefficients revealed that associations between both CDR-SB and ADAS-11 and AD-NeuroScore were significantly stronger than with AHV (p=0.006 for CDR-SB and p=0.024 for ADAS-11). Sub-analyses stratified by baseline diagnosis revealed that AD-NeuroScore and AHV performed the most similarly in participants with MCI, and the most differently in participants with AD (AD-NeuroScore: p<0.01 for ADAS-11 and p<0.05 for CDR-SB, both Holm-Bonferroni corrected, AHV: NS for all).

### 3.2. Longitudinal Validation

#### 3.2.1. Metric Sensitivity to Progression

In the overall baseline experimental cohort, we found that baseline AD-NeuroScore differentiated Diagnosis_stable_ from Diagnosis_decline_ significantly at all timepoints (12, 24, 36, and 48-months; **Table 5**; p<0.001, Holm-Bonferroni corrected, for all). Sub-analyses stratified by baseline diagnosis revealed that AD-NeuroScore’s ability to predict decline was primarily driven by individuals with MCI at baseline (p< 0.001, Holm-Bonferroni corrected, for all). We found no significant effects in CN individuals.

**Table 5.** Relationship Between AD-NeuroScore and Longitudinal Diagnosis Transitions. Sensitivity to diagnostic transition category assessed using pairwise, two-tailed t-tests performed between groups with a stable or declining diagnosis from baseline, at each respective time point. Resulting z-scores, effect sizes (Cohen’s *d*) with 95% confidence intervals (CI)s, and AUC-ROC values with 95% CIs are included. Performance was evaluated both in the overall experimental cohort and in sub-groups based on starting diagnosis (CN or MCI). Results using AHV are included for benchmarking. Significant results from group comparisons are denoted by * to indicate p<0.05, ** to indicate p<0.01, and *** to indicate p<0.001, Holm-Bonferroni corrected.

| Comparison |  | time | Metric | AUC [95% CI] | Cohen's *d* [95% CI] | z-score |
| --- | --- | --- | --- | --- | --- | --- |
| Stable vs. Decline | All | 12 months | ADNS | 0.67 [0.53, 0.80] | 0.64 [0.35, 0.93]*** | -4.35 |
|  |  |  | AHV | 0.65 [0.50, 0.80] | -0.53 [-0.81, -0.24]** | -3.57 |
|  |  | 24 months | ADNS | 0.71 [0.59, 0.82] | 0.84 [0.61, 1.08]*** | -7.00 |
|  |  |  | AHV | 0.71 [0.61, 0.82] | -0.79 [-1.02, -0.55]*** | -6.58 |
|  |  | 36 months | ADNS | 0.69 [0.53, 0.85] | 0.72 [0.46, 0.98]*** | -5.42 |
|  |  |  | AHV | 0.72 [0.60, 0.85] | -0.79 [-1.05, -0.52]*** | -5.88 |
|  |  | 48 months | ADNS | 0.67 [0.52, 0.83] | 0.74 [0.47, 1.01]*** | -5.36 |
|  |  |  | AHV | 0.72 [0.62, 0.83] | -0.79 [-1.06, -0.51]*** | -5.66 |
|  | CN | 12 months | ADNS | 0.35 [0.08, 0.62] | -0.06 [-0.70, 0.59] | -0.17 |
|  |  |  | AHV | 0.72 [0.35, 1.09] | -0.59 [-1.23, 0.06] | -1.79 |
|  |  | 24 months | ADNS | 0.64 [0.41, 0.88] | 0.55 [0.08, 1.01] | -2.32 |
|  |  |  | AHV | 0.74 [0.53, 0.95] | -0.79 [-1.26, -0.33]** | -3.34 |
|  |  | 36 months | ADNS | 0.59 [0.21, 0.97] | 0.34 [-0.36, 1.05] | -0.98 |
|  |  |  | AHV | 0.78 [0.46, 1.09] | -0.84 [-1.57, -0.11] | -2.30 |
|  |  | 48 months | ADNS | 0.44 [0.14, 0.75] | 0.11 [-0.44, 0.65] | -0.39 |
|  |  |  | AHV | 0.72 [0.46, 0.98] | -0.65 [-1.20, -0.10] | -2.34 |
|  | MCI | 12 months | ADNS | 0.77 [0.65, 0.90] | 1.08 [0.75, 1.42]*** | -6.39 |
|  |  |  | AHV | 0.68 [0.51, 0.85] | -0.69 [-1.02, -0.36]*** | -4.14 |
|  |  | 24 months | ADNS | 0.76 [0.64, 0.88] | 1.05 [0.77, 1.34]*** | -7.36 |
|  |  |  | AHV | 0.73 [0.59, 0.86] | -0.85 [-1.12, -0.57]*** | -5.99 |
|  |  | 36 months | ADNS | 0.73 [0.62, 0.84] | 0.87 [0.59, 1.16]*** | -5.99 |
|  |  |  | AHV | 0.73 [0.56, 0.90] | -0.86 [-1.14, -0.57]*** | -5.87 |
|  |  | 48 months | ADNS | 0.73 [0.57, 0.89] | 0.78 [0.46, 1.10]*** | -4.83 |
|  |  |  | AHV | 0.69 [0.51, 0.88] | -0.76 [-1.08, -0.44]*** | -4.71 |

We found that AD-NeuroScore performed as well as AHV in the overall experimental cohort (AHV: p<0.01 at 12-months, p<0.001 at all successive sessions, Holm-Bonferroni corrected). While there were no significant differences between AD-NeuroScore and AHV, qualitatively, AD-NeuroScore tended to do modestly better than AHV at differentiating Diagnosis_stable,_ from Diagnosis_decline_ participants at 12- and 24-months, and slightly worse at 36- and 48-months. Sub-analysis by baseline diagnosis revealed similar patterns; AD-NeuroScore performed equivalently to AHV in individuals with MCI. However, AHV demonstrated somewhat

#### 3.2.2. Longitudinal Association with Disease Severity (MMSE, ADAS-11, and CDR-SB)

In the overall longitudinal cohort, we found that baseline AD-NeuroScore was also significantly associated with the change in disease severity scores (**Table 4**; p <0.001, Holm-Bonferroni corrected, for all), as measured by the change in MMSE, ADAS-11, and CDR-SB scores from baseline to each time point (12, 24, 36, and 48-months). Sub-analysis by baseline diagnosis revealed that AD-NeuroScore’s association with the change in disease severity scores was also primarily driven by individuals with MCI at baseline. We found significant associations between AD-NeuroScore and the change in MMSE, ADAS-11, and CDR-SB scores in participants with MCI at all time points (p<0.001, Holm-Bonferroni corrected for all). Conversely, we found no significant association with the change in disease severity in AD individuals, and only two significant associations in CN individuals (CDR-SB at 12- and 24-months; p<0.05, Holm-Bonferroni corrected, for both).

AD-NeuroScore generally performed as well or better than our benchmark, AHV, in this longitudinal validation (**Table 4**). As with AD-NeuroScore, correlations between AHV at baseline and change in disease severity scores were significant in the overall longitudinal cohort (**Table 4**; p<0.001, Holm-Bonferroni corrected, for all timepoints). Results from z-tests conducted using Fisher z-transformed correlation coefficients revealed that correlations between AD-NeuroScore and 24-month change in ADAS-11 were significantly stronger than with AHV (p=0.003, Holm-Bonferroni corrected). In the MCI group, AD-NeuroScore performed as well or somewhat better than AHV (AD-NeuroScore: p<0.001 for all; AHV: p<0.05 for MMSE across all time points, p<0.01 for ADAS-11 at 12-months, and p<0.001 for all other scores and time points, all Holm-Bonferroni corrected).

In the AD group, neither AD-NeuroScore nor AHV were significantly associated with change in disease severity scores. The AD sub-analysis was limited by sample size longitudinally. In the CN group, both AD-NeuroScore and AHV had generally weak correlations, none of which were significant aside from AD-NeuroScore and CDR-SB at 12- and 24-months (both p<0.05, Holm-Bonferroni corrected).

### 3.3. Alternative Atlas Analysis

#### 3.3.1 Baseline and Longitudinal Validation

Results from analyses repeated using the Neuroreader® ROI-based AD-NeuroScore were similar to those obtained with the Freesurfer-based AD-NeuroScore, including all cross-sectional and longitudinal analyses. Benchmarking the pseudo-Neuroreader® ROI-based AD-NeuroScore against AHV also produced results analogous to those of the original analysis (**Supplementary Tables S5-S7)**.

## 4. Discussion

We developed a Euclidean inspired sMRI-based distance metric, AD-NeuroScore, which was significantly associated with diagnosis (CN, MCI, and AD) and disease severity (MMSE, ADAS-11 and CDR-SB scores) at baseline. AD-NeuroScore performed as well as AHV, the most commonly used sMRI measure of AD-related neurodegeneration, and demonstrated comparably strong correlations with disease severity scores. AD-NeuroScore was also significantly associated with changes in the three disease severity scores over a relatively long follow-up period of 48 months. We found that these associations were largely driven by the MCI group. It is possible that the limited sample size of CN participants and patients with AD with longitudinal scores available resulted in the CN and AD sub-analyses being underpowered to detect a relationship.

Moreover, AD-NeuroScore was also able to differentiate between participants who declined in cognitive status (from CN to MCI or AD; from MCI to AD) and those who remained stable in longitudinal analyses; this was also true of AHV. Qualitatively, AD-NeuroScore tended to perform modestly better than AHV at earlier time points (12- and 24-months) and slightly worse at later sessions (36- and 48-months). Stratifying this analysis by starting diagnosis revealed that the MCI patient population was primarily driving these results as well. This finding was likely the result of two major factors: the combined sample of stable and declining patients at each time point was always much larger for the patients with a baseline MCI diagnosis and the greatest and most consistent changes in neuropsychological scale scores across sessions would be expected to occur in the MCI sub-population. Because patients more frequently seek monitoring and intervention after the onset of cognitive deficits (i.e., with MCI) rather than prior (i.e., CN), the strength of our metric in this patient population is of importance. Similarly, in clinical trials aimed at assessing the efficacy of interventions in slowing AD progression, a measure that is optimally sensitive in patients with MCI is particularly well-suited for usage.

A particularly attractive feature of AD-NeuroScore is the ease with which it could be integrated into current clinical and research workflows. One of the challenges in the full clinical implementation of AI applications is the difficulty integrating these into daily workflows (Hwang and Park, 2020). Adding AD-NeuroScore to existing, clinically implemented regional brain volume reports, and calculating AD-NeuroScore based on the individual regional volumes obtained in research settings would be straight-forward. Further, by replacing different anatomical ROIs with a single measure as an endpoint, AD-NeuroScore could enhance the sensitivity of many studies by reducing the number of statistical tests, while meaningfully encapsulating as much or more information. Future directions of this work include creating validated AD-NeuroScore norms and diagnostic cutoffs to aid in integration with clinical workflows.

An unexpected result of the present study, inconsistent with previous literature, is that both the left and right lateral ventricular volumes were not retained during ROI selection, despite the frequent observation of ventricular dilation in patients with AD (Attier-Zmudka et al., 2019; Ott et al., 2010). Absolute ventricular enlargement has proven sufficient to discriminate between MCI patients who progressed to AD or remained stable over a 6-month interval in a smaller ADNI cohort (n=504) and, consequently, has been proposed as a potential biomarker for disease progression (Sean M. Nestor et al., 2008). However, at baseline, in line with previous cross-sectional volumetric studies, a large overlap was found between the total ventricular volumes of pathological groups and controls (Giesel et al., n.d.; Sean M. Nestor et al., 2008; Schott et al., 2005). Thus, one potential explanation may be that absolute ventricular change is a more sensitive measure of AD-pathology than total ventricular volume. Given that our investigation only examined regional volumes at cross-section and employed a conservative correction to adjust for the 84 brain regions tested during ROI selection, this discrepancy may be plausibly attributed to differences in study design.

It is also important to address the distinctions between AD-NeuroScore and other biomarkers that have been developed and studied in the literature. Many of the ML-based metrics successfully classify participants as AD or not, using similar input features (perhaps from different data sets) and slightly different ML algorithms. The STructural Abnormality iNDex (STAND)-score uses a support vector machine classifier which takes in sMRI normalized input features such as GM, WM, and CSF tissue densities extracted from SPM5 tissue segmentation as well as demographic information and assigns a numerical value to classify a patient as CN or AD based on a 280 sample (140 each) training set from the Alzheimer’s disease Patient Registry (Vemuri et al., 2009b, n.d.). AD Pattern Similarity (AD-PS) uses the same sMRI input features (GM, WM, and CSF tissue maps) derived from the ADNI data set (Casanova et al., 2013) and high dimensional regularized logistic regression to classify a patient as CN or AD. AD-PS and AD-NeuroScore perform similarly (with significantly overlapping AUC-ROC confidence intervals); while AD-PS was also shown to correlate negatively with cognitive scores obtained through clinic visits and telephone interviews, lack of details regarding the correlations makes direct comparison with AD-NeuroScore difficult. Another ML-based metric, Subtype and Stage Inference (SuStaIn) (Young et al., 2014), primarily subtypes individuals based on their likelihood to stabilize or decline (Fonteijn et al., 2012) and excels at differentiating AD from CN. AD-NeuroScore performs at least as well as SuStaIn at subtyping MCI patients (AUC=0.72 for SuStaIn; AUC=0.77 for AD-NeuroScore) (Archetti et al., 2021). SuStaIn utilizes multi-modal sporadic disease data rather than a single data source, making it more complicated than AD-NeuroScore to integrate into clinical workflows, potentially a barrier to full clinical implementation.

Amongst the more interpretable ML models, MRDATS and SPARE-AD (Davatzikos et al., 2009; Popuri et al., 2020b) deserve mention. MRDATS uses w-corrected brain ROI volumes in an ensemble-learning algorithm to arrive at a score between 0 and 1 to indicate CN to dementia progression (Popuri et al., 2020b). Using a threshold of 0.5, MRDATS performed well at binary classification of CN and dementia, however this was assessed only in the training set. As with SuStaIn, MRDATS was developed primarily to differentiate stable from declining patients. MRDATS performed equivalently to AD-NeuroScore in distinguishing stable and progressive MCI groups (0–3-year AUC=0.75 for MRDATS and AUC=0.75 for AD-NeuroScore). The SPARE-AD (Davatzikos et al., 2009) index uses high-dimensional pattern classification of volumetric atrophy and rate of change of SPARE-AD was highly predictive of cognitive status. A future direction of this work is to determine if utilizing rate of change of AD-NeuroScore (versus a single time point) improves its performance. Compared to MRDATS and SPARE-AD, AD-NeuroScore can be calculated in a computationally simple way using FDA-approved, clinically implemented, automated ROI-metrics such as Neuroreader®.

The most similar metric to AD-NeuroScore is RVI or the ENIGMA Dot Product (Kochunov et al., 2022), which also uses the concept of weighing a multivariate sum by a metric describing the effect of a group status with harmonized phenotypes and performs well as a biomarker in several neuropsychiatric conditions. There are many notable differences between RVI and AD-NeuroScore, the first of which pertains to the phenotypes used within the metric. AD-NeuroScore exclusively uses regional volumes while RVI uses several brain-derived metrics including gray matter thicknesses and fractional anisotropy values, which are not presently part of popular clinical workflows. Moreover, AD-NeuroScore retains both hemispheric volumes compared to RVI’s averaging of the hemispheres. AD-NeuroScore additionally harmonizes with respect to scanner model, improving its utility in multi-scanner circumstances, which are common in large cohort data sets and in clinical practice.

Most of the important differences arise in the construction of AD-NeuroScore. The core concept of the metric is the Euclidean distance, where each harmonized volume is subtracted from the corresponding normal template harmonized volume. Unlike RVI, it does not take the absolute value of the transformed residual after harmonization. For this reason, each multivariate term in the sum is squared to account for possible cancellations of differences and to emphasize differences that are greater while minimizing smaller ones. Moreover, AD-NeuroScore quantifies the difference between groups, weighs each term in the sum with a z-score rather than effect size and does not normalize by the number of terms. These differences could explain why AD-NeuroScore better distinguishes CN and AD groups from the ADNI database than RVI, with statistically significant differences in their Cohen’s d values (p<0.001) (Kochunov et al., 2021).

An important advantage of AD-NeuroScore is its potential to accurately monitor disease advancement and be compared across multiple sites by using standard ranges to generate probabilities of diagnosis or decline. Similarly, ranges of AD-NeuroScore that are standard for different groups could be used to aid early differential diagnosis of common neurodegenerative diseases that cause dementia, such as vascular dementia, dementia with Lewy bodies, and frontotemporal dementia, which demonstrate unique patterns of atrophy that may be difficult to assess by AHV and other standard approaches. In support of exploring this functionality further is previous literature which has found that sMRI alone can be used to differentiate between these four dementia-causing neurodegenerative diseases groups and healthy controls with a high accuracy (Koikkalainen et al., 2016). A future direction of this research is to determine if AD-NeuroScore can aid in the differential diagnosis of dementia.

While the findings of this work encourage further investigation, there are several limitations that should be carefully considered and addressed in successive studies. First, the sample of patients with AD was much smaller than the CN and MCI cohorts across all sessions, with this number falling across successive visits. The limited number of participants across all diagnostic groups with scores available on the MMSE, CDR-SB, and ADAS-11 also dwindled longitudinally, rendering some of the later tests for the CN and AD groups particularly underpowered. Additionally, the AD subset was confined to patients with mild AD dementia, as defined in the ADNI protocol, which leaves more progressed patients with AD understudied with regards to our metric. Repeating this study in a real-world clinical sample that better represents the AD patient class at various stages of disease advancement would solidify the findings presented in this study.

## Data Availability

Full, open access to all de-identified ADNI imaging and clinical data is publicly and freely available to individuals who register with the ADNI and agree to the conditions in the “ADNI Data Use Agreement,” upon approval of a request that includes the proposed analysis and the named lead investigator (contact via http://adni.loni.usc.edu/data-samples/access-data/). Additional details about the ADNI data acquisition and sharing policies can be found at http://adni.loni.usc.edu/wp-content/uploads/how_to_apply/ADNI_DSP_Policy.pdf.
The code supporting the findings of this study is openly available in [repository name: "AD-NeuroScore"] at https://github.com/jbramen/AD-NeuroScore. The Python programming language (version 3.9) was used for all analyses (Python Software Foundation, https://www.python.org/).

https://github.com/jbramen/AD-NeuroScore

http://adni.loni.usc.edu/data-samples/access-data/

## Abbreviations

(AD): Alzheimer’s Disease
(ADAS-11): Alzheimer’s Disease Assessment Scale-Cognitive Subscale
(aMCI): Amnestic Mild Cognitive Impairment
(AUC-ROC): Area Under the Receiver Operator Characteristic Curve
(CSF): Cerebrospinal Fluid
(CDR-SB): Clinical Dementia Rating Scale Sum of Boxes
(GM): Grey Matter
(ML): Machine Learning
(MTL): Medial Temporal Lobe
(MCI): Mild Cognitive Impairment
(MMSE): Mini-Mental State Exam
(PHI): Protected Health Information
(ROC): Receiver Operator Characteristic
(ROI): Region of Interest
(SD): Standard Deviation
(sMRI): Structural Magnetic Resonance imaging
(WM): White Matter

## Acknowledgements

We thank the participants and families volunteering for this research. We thank the Pacific Neuroscience Institute and Foundation staff and leadership, including the CEO and Founder, Dan Kelly, MD, Vice President, Melissa Coleman, Director of Research Administration and Operations Melanie Lampa, Director of Marketing, Zara Jethani, Executive Assistant, Danielle Wozniak, and Accounting Manager, Bersabel Belay and the Pacific Brain Health Clinic’s Practice Manager, Brenda Smith for their support. This work is supported by the Pacific Neuroscience Institute Foundation, including the generous support of the Barbara and John McLoughlin Family as well as the Cary and Will Singleton Family. Data collection and sharing for this project was funded by the Alzheimer’s Disease Neuroimaging Initiative (ADNI) (National Institutes of Health Grant U01 AG024904) and DOD ADNI (Department of Defense award number W81XWH-12-2-0012). ADNI is funded by the National Institute on Aging, the National Institute of Biomedical Imaging and Bioengineering, and through generous contributions from the following: AbbVie, Alzheimer’s Association; Alzheimer’s Drug Discovery Foundation; Araclon Biotech; BioClinica, Inc.; Biogen; Bristol-Myers Squibb Company; CereSpir, Inc.; Cogstate; Eisai Inc.; Elan Pharmaceuticals, Inc.; Eli Lilly and Company; EuroImmun; F. Hoffmann-La Roche Ltd and its affiliated company Genentech, Inc.; Fujirebio; GE Healthcare; IXICO Ltd.; Janssen Alzheimer Immunotherapy Research & Development, LLC.; Johnson & Johnson Pharmaceutical Research & Development LLC.; Lumosity; Lundbeck; Merck & Co., Inc.; Meso Scale Diagnostics, LLC.; NeuroRx Research; Neurotrack Technologies; Novartis Pharmaceuticals Corporation; Pfizer Inc.; Piramal Imaging; Servier; Takeda Pharmaceutical Company; and Transition Therapeutics. The Canadian Institutes of Health Research is providing funds to support ADNI clinical sites in Canada. Private sector contributions are facilitated by the Foundation for the National Institutes of Health (www.fnih.org). The grantee organization is the Northern California Institute for Research and Education, and the study is coordinated by the Alzheimer’s Therapeutic Research Institute at the University of Southern California. ADNI data are disseminated by the Laboratory for Neuro Imaging at the University of Southern California.

---

**Supplemental Table 1.** Baseline Assessment of all Distance Metrics based on Sensitivity to Diagnosis. Assessment of all distance metrics, benchmarked against AHV, based on metric sensitivity to diagnosis at baseline, operationalized as pairwise, two-tailed t-tests performed for each possible diagnostic group comparison, z-scores, effect sizes (Cohen’s *d*) with 95% CIs, and AUC values with 95% CIs. AD-NeuroScore (ADNS) is used to refer to the Z-Weighted Euclidean (ZWE) distance function. The Fréchet and Hausdorff distance functions were found to be the same in one dimension. Significant group comparisons are denoted by * to indicate p<0.05, ** to indicate p<0.01, and *** to indicate p<0.001.

| Group Comparison | Metric | AUC [95% CI] | Cohen's *d* [95% CI] | z-score |
| --- | --- | --- | --- | --- |
| CN vs MCI | ADNS | 0.66 [0.59, 0.73] | -0.56 [-0.59, -0.53]*** | -7.46 |
|  | Euclidean | 0.64 [0.55, 0.73] | -0.47 [-0.50, -0.44]*** | -6.31 |
|  | 1-D Fréchet | 0.65 [0.57, 0.73] | -0.49 [-0.52, -0.46]*** | -6.57 |
|  | N-D Fréchet | 0.58 [0.49, 0.66] | -0.33 [-0.35, -0.30]*** | -4.41 |
|  | 1-D Hausdorff | 0.63 [0.55, 0.71] | -0.49 [-0.52, -0.46]*** | -6.57 |
|  | N-D Hausdorff | 0.56 [0.49, 0.63] | -0.22 [-0.24, -0.19]** | -2.95 |
|  | AHV | 0.65 [0.58, 0.72] | 0.55 [0.52, 0.58]*** | -7.27 |
| CN vs AD | ADNS | 0.91 [0.86, 0.96] | -2.06 [-2.24, -1.88]*** | -16.37 |
|  | Euclidean | 0.85 [0.78, 0.92] | -1.44 [-1.60, -1.28]*** | -12.32 |
|  | 1-D Fréchet | 0.85 [0.78, 0.93] | -1.57 [-1.73, -1.40]*** | -13.23 |
|  | N-D Fréchet | 0.80 [0.71, 0.88] | -1.28 [-1.44, -1.12]*** | -11.18 |
|  | 1-D Hausdorff | 0.86 [0.79, 0.93] | -1.57 [-1.73, -1.40]*** | -13.23 |
|  | N-D Hausdorff | 0.73 [0.62, 0.84] | -0.93 [-1.08, -0.78]*** | -8.40 |
|  | AHV | 0.88 [0.81, 0.95] | 1.72 [1.55, 1.89]*** | -14.27 |
| MCI vs AD | ADNS | 0.80 [0.71, 0.89] | -1.16 [-1.19, -1.14]*** | -11.23 |
|  | Euclidean | 0.74 [0.64, 0.85] | -1.02 [-1.05, -1.00]*** | -9.98 |
|  | 1-D Fréchet | 0.74 [0.64, 0.84] | -1.02 [-1.05, -1.00]*** | -9.99 |
|  | N-D Fréchet | 0.73 [0.63, 0.83] | -0.85 [-0.87, -0.83]*** | -8.41 |
|  | 1-D Hausdorff | 0.74 [0.63, 0.84] | -1.02 [-1.05, -1.00]*** | -9.99 |
|  | N-D Hausdorff | 0.64 [0.53, 0.75] | -0.68 [-0.71, -0.66]*** | -6.82 |
|  | AHV | 0.76 [0.67, 0.84] | 0.92 [0.90, 0.94]*** | -9.04 |

**Supplemental Table 2.**
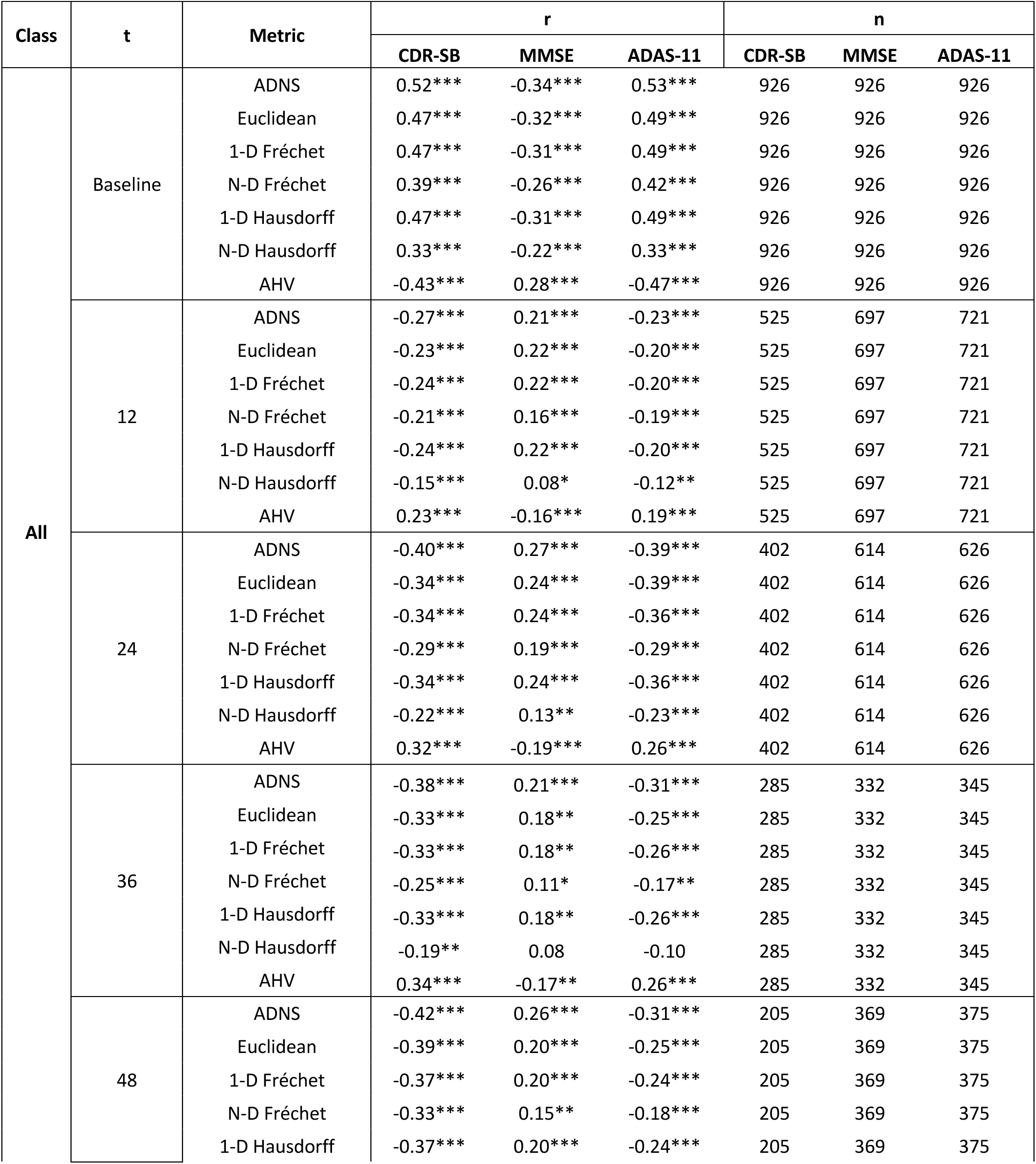
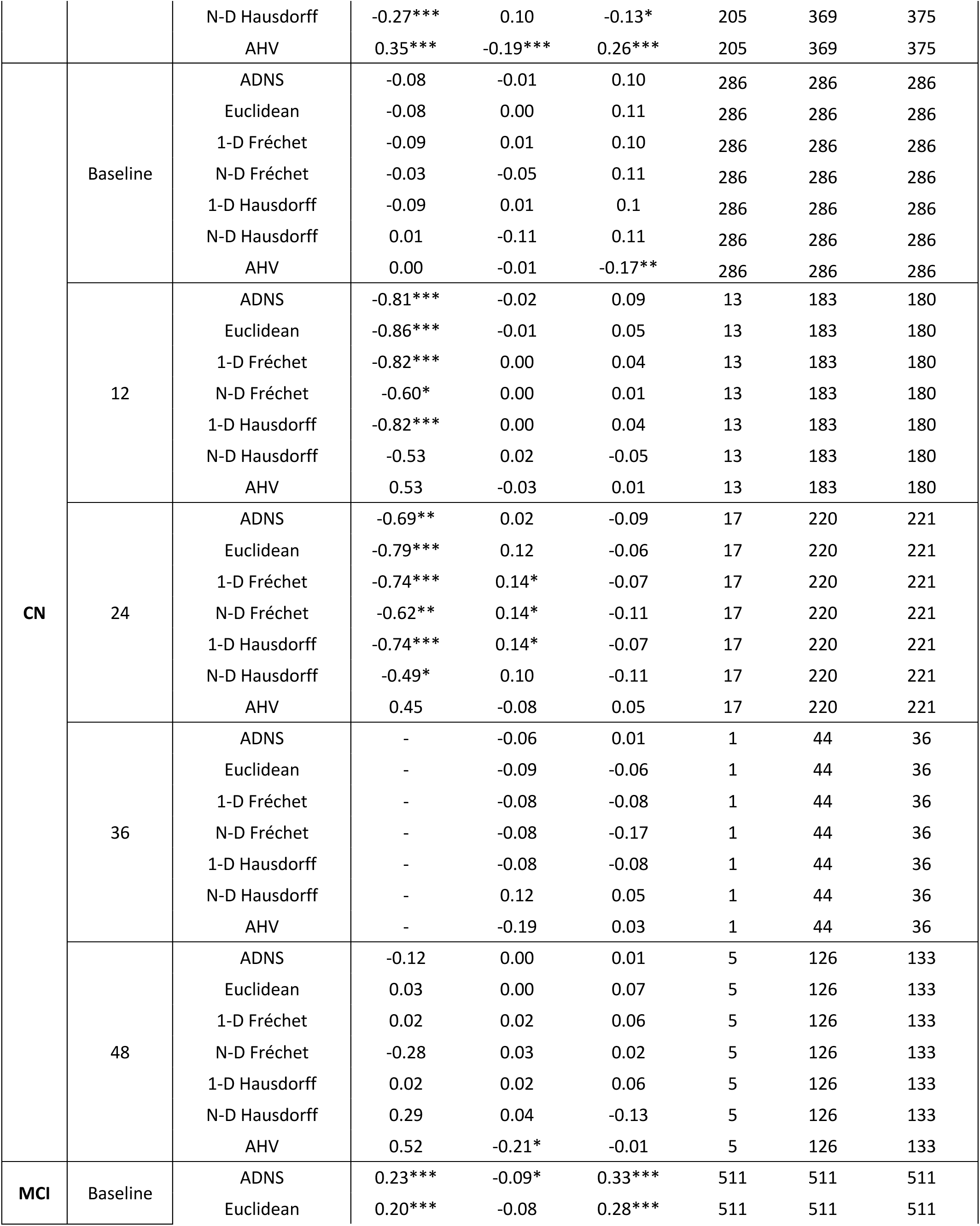
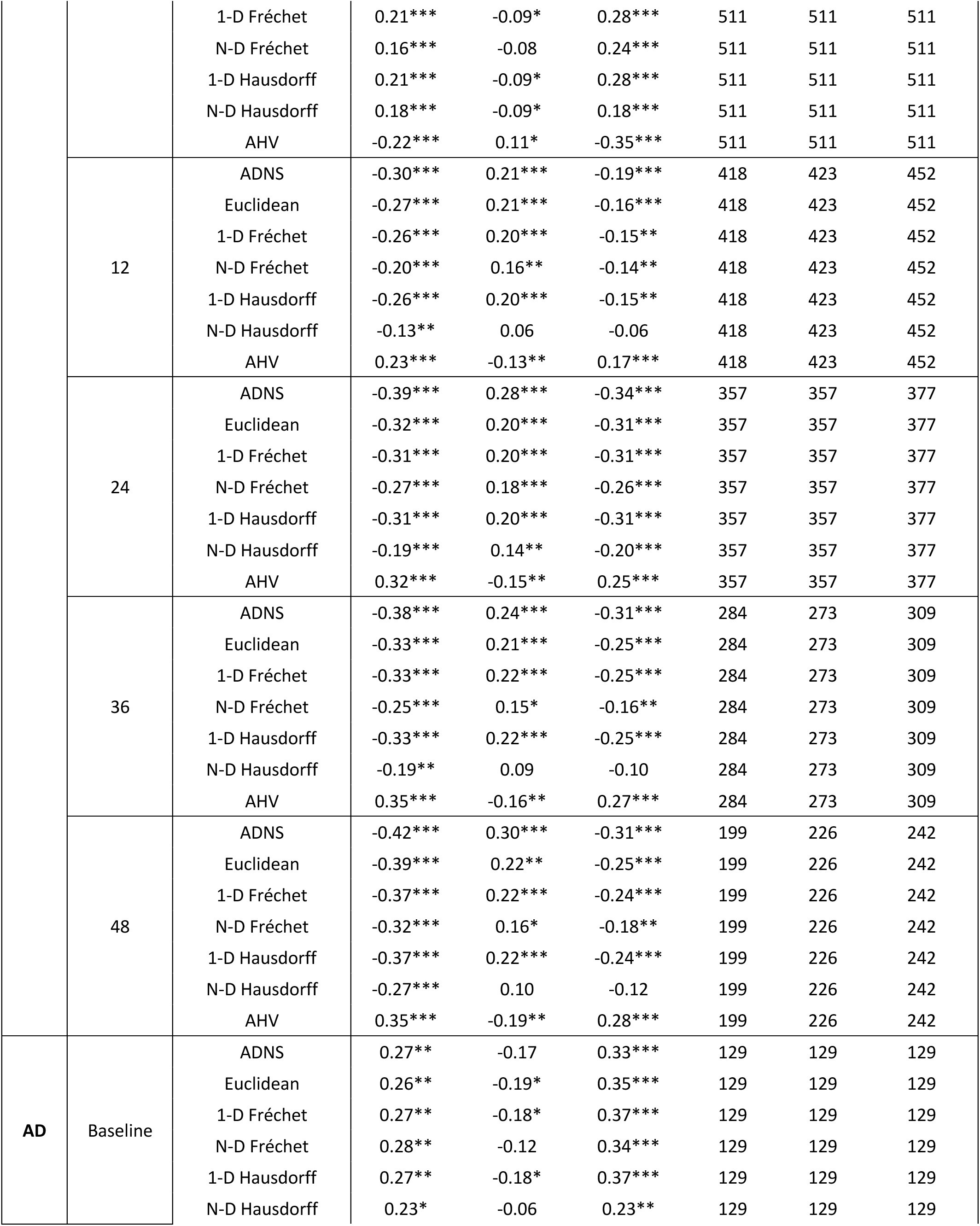
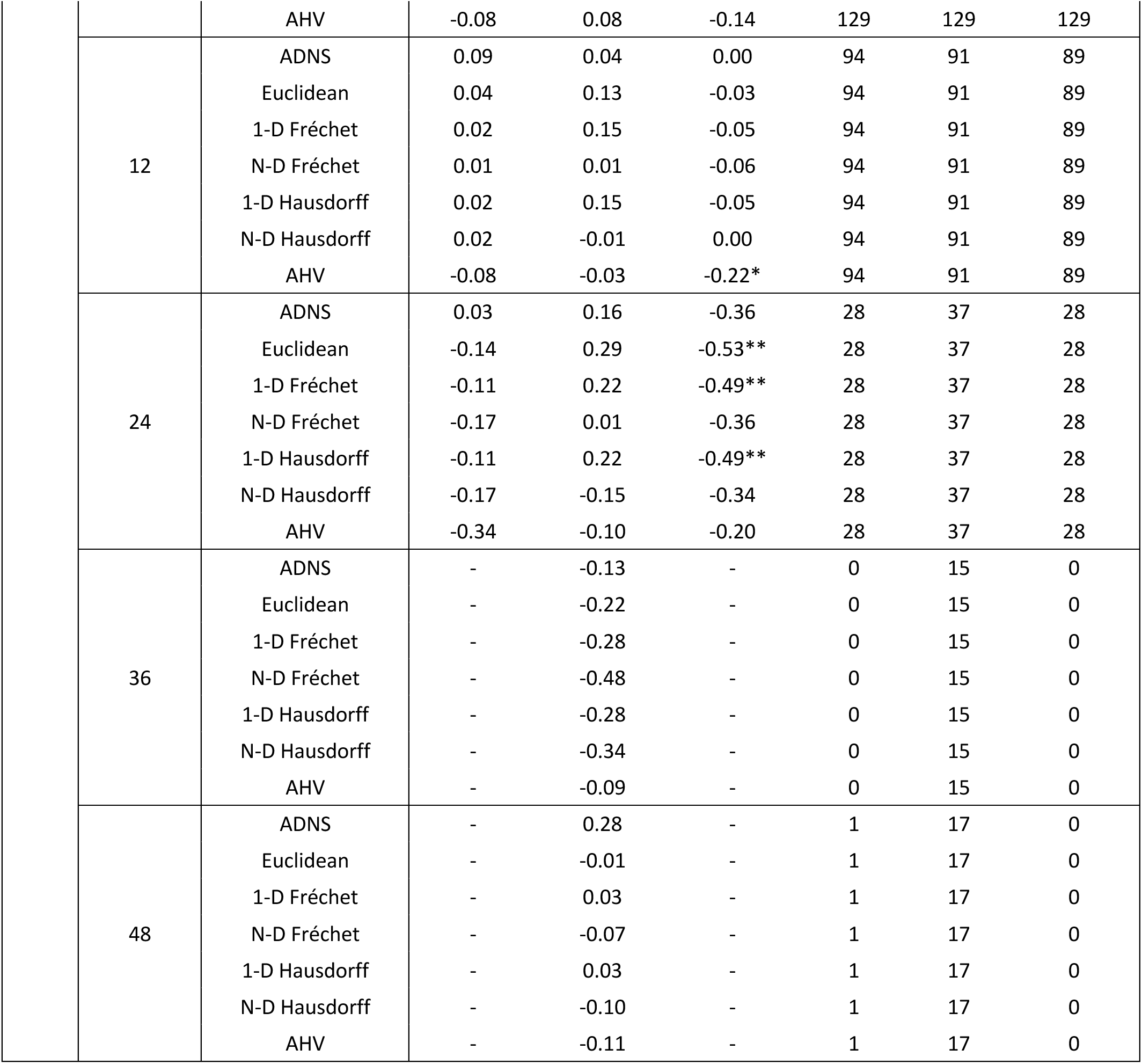
Linear Regression Results All Distance Metrics. Assessment of all distance metrics, benchmarked against AHV, on the basis of metric association with disease severity, operationalized as MMSE, ADAS-11, and CDR-SB scores at baseline or their change from baseline at each time point, tested with linear regression in the overall experimental cohort and in each diagnostic sub-group. ADNS is used here to refer to the Z-Weighted Euclidean (ZWE) distance function. The Fréchet and Hausdorff distance functions were found to be the same in one dimension. Significant group comparisons are denoted by * to indicate p<0.05, ** to indicate p<0.01, and *** to indicate p<0.001.

**Supplemental Table 3.**
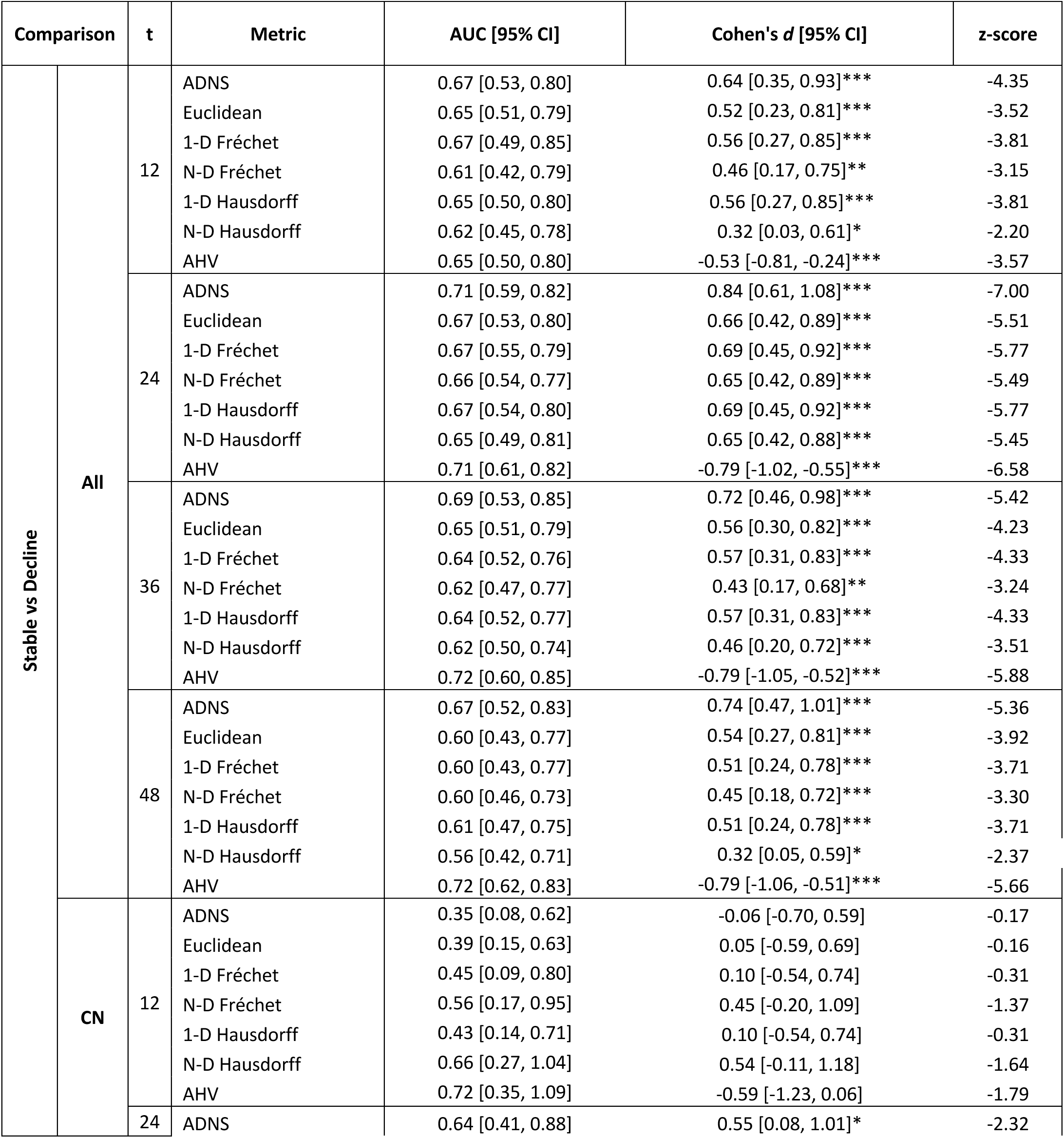
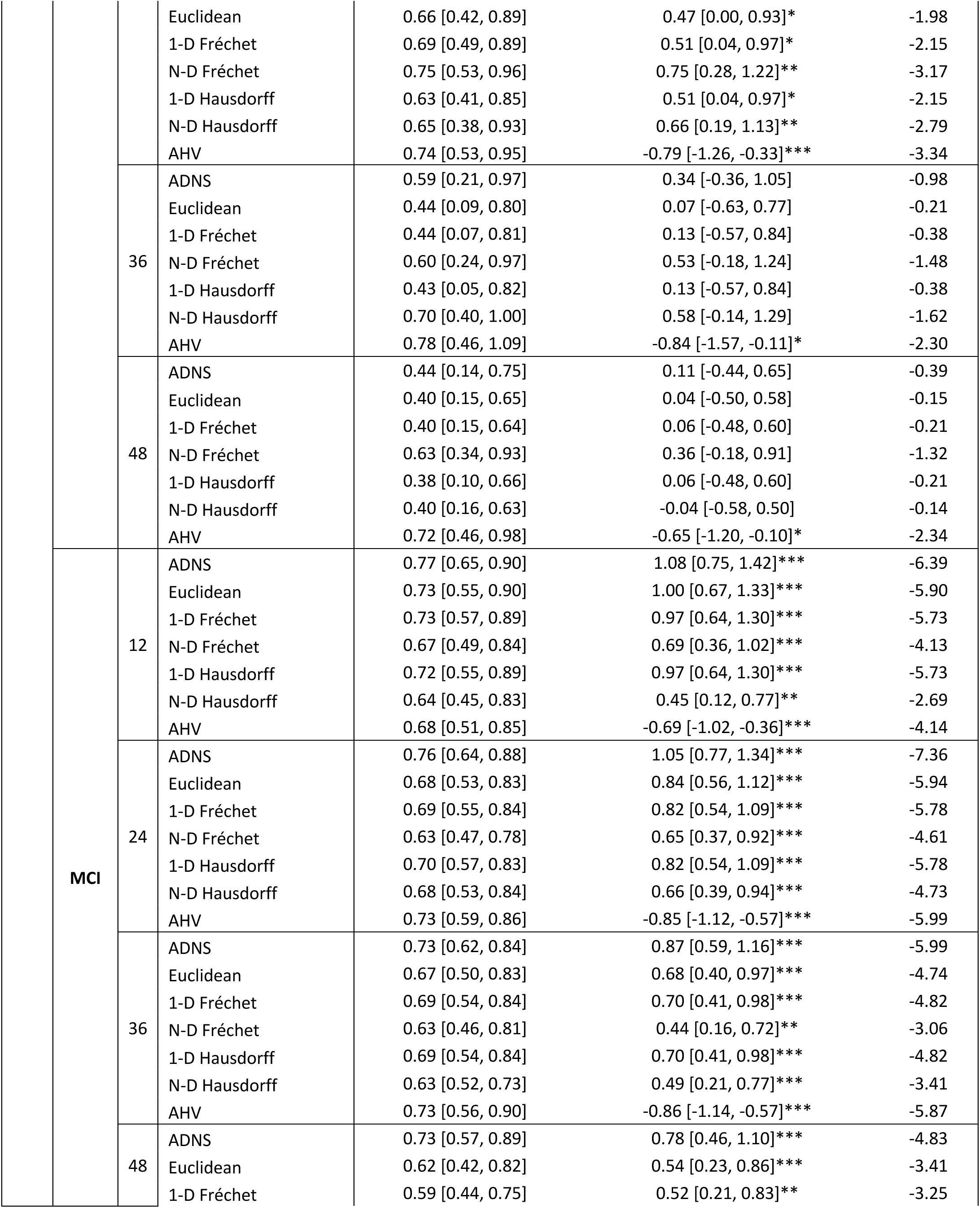
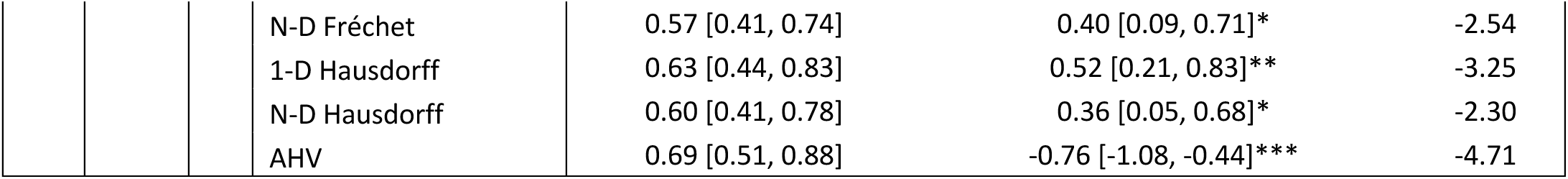
Longitudinal Assessment of all Distance Metrics based on Diagnosis Transitions. Assessment of all distance metrics, benchmarked against AHV, on the basis of metric ability to predict progression, operationalized as two-tailed t-tests performed for each DTC comparison, z-scores, effect sizes (Cohen’s *d*) with 95% CIs, and AUC-ROC with 95% CIs. Evaluation was performed in the full experimental cohort and in sub-groups based on starting diagnosis. ADNS refers to the Z-Weighted Euclidean (ZWE) distance function. The Fréchet and Hausdorff distance functions were found to be the same in one dimension. Significant group comparisons are denoted by * to indicate p<0.05, ** to indicate p<0.01, and *** to indicate p<0.001

| Comparison |  | t | Metric | AUC [95% CI] | Cohen's *d* [95% CI] | z-score |
| --- | --- | --- | --- | --- | --- | --- |
| Stable vs Decline | All | 12 | ADNS | 0.67 [0.53, 0.80] | 0.64 [0.35, 0.93]*** | -4.35 |
|  |  |  | Euclidean | 0.65 [0.51, 0.79] | 0.52 [0.23, 0.81]*** | -3.52 |
|  |  |  | 1-D Fréchet | 0.67 [0.49, 0.85] | 0.56 [0.27, 0.85]*** | -3.81 |
|  |  |  | N-D Fréchet | 0.61 [0.42, 0.79] | 0.46 [0.17, 0.75]** | -3.15 |
|  |  |  | 1-D Hausdorff | 0.65 [0.50, 0.80] | 0.56 [0.27, 0.85]*** | -3.81 |
|  |  |  | N-D Hausdorff | 0.62 [0.45, 0.78] | 0.32 [0.03, 0.61]* | -2.20 |
|  |  |  | AHV | 0.65 [0.50, 0.80] | -0.53 [-0.81, -0.24]*** | -3.57 |
|  |  | 24 | ADNS | 0.71 [0.59, 0.82] | 0.84 [0.61, 1.08]*** | -7.00 |
|  |  |  | Euclidean | 0.67 [0.53, 0.80] | 0.66 [0.42, 0.89]*** | -5.51 |
|  |  |  | 1-D Fréchet | 0.67 [0.55, 0.79] | 0.69 [0.45, 0.92]*** | -5.77 |
|  |  |  | N-D Fréchet | 0.66 [0.54, 0.77] | 0.65 [0.42, 0.89]*** | -5.49 |
|  |  |  | 1-D Hausdorff | 0.67 [0.54, 0.80] | 0.69 [0.45, 0.92]*** | -5.77 |
|  |  |  | N-D Hausdorff | 0.65 [0.49, 0.81] | 0.65 [0.42, 0.88]*** | -5.45 |
|  |  |  | AHV | 0.71 [0.61, 0.82] | -0.79 [-1.02, -0.55]*** | -6.58 |
|  |  | 36 | ADNS | 0.69 [0.53, 0.85] | 0.72 [0.46, 0.98]*** | -5.42 |
|  |  |  | Euclidean | 0.65 [0.51, 0.79] | 0.56 [0.30, 0.82]*** | -4.23 |
|  |  |  | 1-D Fréchet | 0.64 [0.52, 0.76] | 0.57 [0.31, 0.83]*** | -4.33 |
|  |  |  | N-D Fréchet | 0.62 [0.47, 0.77] | 0.43 [0.17, 0.68]** | -3.24 |
|  |  |  | 1-D Hausdorff | 0.64 [0.52, 0.77] | 0.57 [0.31, 0.83]*** | -4.33 |
|  |  |  | N-D Hausdorff | 0.62 [0.50, 0.74] | 0.46 [0.20, 0.72]*** | -3.51 |
|  |  |  | AHV | 0.72 [0.60, 0.85] | -0.79 [-1.05, -0.52]*** | -5.88 |
|  |  | 48 | ADNS | 0.67 [0.52, 0.83] | 0.74 [0.47, 1.01]*** | -5.36 |
|  |  |  | Euclidean | 0.60 [0.43, 0.77] | 0.54 [0.27, 0.81]*** | -3.92 |
|  |  |  | 1-D Fréchet | 0.60 [0.43, 0.77] | 0.51 [0.24, 0.78]*** | -3.71 |
|  |  |  | N-D Fréchet | 0.60 [0.46, 0.73] | 0.45 [0.18, 0.72]*** | -3.30 |
|  |  |  | 1-D Hausdorff | 0.61 [0.47, 0.75] | 0.51 [0.24, 0.78]*** | -3.71 |
|  |  |  | N-D Hausdorff | 0.56 [0.42, 0.71] | 0.32 [0.05, 0.59]* | -2.37 |
|  |  |  | AHV | 0.72 [0.62, 0.83] | -0.79 [-1.06, -0.51]*** | -5.66 |
|  | CN | 12 | ADNS | 0.35 [0.08, 0.62] | -0.06 [-0.70, 0.59] | -0.17 |
|  |  |  | Euclidean | 0.39 [0.15, 0.63] | 0.05 [-0.59, 0.69] | -0.16 |
|  |  |  | 1-D Fréchet | 0.45 [0.09, 0.80] | 0.10 [-0.54, 0.74] | -0.31 |
|  |  |  | N-D Fréchet | 0.56 [0.17, 0.95] | 0.45 [-0.20, 1.09] | -1.37 |
|  |  |  | 1-D Hausdorff | 0.43 [0.14, 0.71] | 0.10 [-0.54, 0.74] | -0.31 |
|  |  |  | N-D Hausdorff | 0.66 [0.27, 1.04] | 0.54 [-0.11, 1.18] | -1.64 |
|  |  |  | AHV | 0.72 [0.35, 1.09] | -0.59 [-1.23, 0.06] | -1.79 |
|  |  | 24 | ADNS | 0.64 [0.41, 0.88] | 0.55 [0.08, 1.01]* | -2.32 |
|  |  |  | Euclidean | 0.66 [0.42, 0.89] | 0.47 [0.00, 0.93]* | -1.98 |
|  |  |  | 1-D Fréchet | 0.69 [0.49, 0.89] | 0.51 [0.04, 0.97]* | -2.15 |
|  |  |  | N-D Fréchet | 0.75 [0.53, 0.96] | 0.75 [0.28, 1.22]** | -3.17 |
|  |  |  | 1-D Hausdorff | 0.63 [0.41, 0.85] | 0.51 [0.04, 0.97]* | -2.15 |
|  |  |  | N-D Hausdorff | 0.65 [0.38, 0.93] | 0.66 [0.19, 1.13]** | -2.79 |
|  |  |  | AHV | 0.74 [0.53, 0.95] | -0.79 [-1.26, -0.33]*** | -3.34 |
|  |  | 36 | ADNS | 0.59 [0.21, 0.97] | 0.34 [-0.36, 1.05] | -0.98 |
|  |  |  | Euclidean | 0.44 [0.09, 0.80] | 0.07 [-0.63, 0.77] | -0.21 |
|  |  |  | 1-D Fréchet | 0.44 [0.07, 0.81] | 0.13 [-0.57, 0.84] | -0.38 |
|  |  |  | N-D Fréchet | 0.60 [0.24, 0.97] | 0.53 [-0.18, 1.24] | -1.48 |
|  |  |  | 1-D Hausdorff | 0.43 [0.05, 0.82] | 0.13 [-0.57, 0.84] | -0.38 |
|  |  |  | N-D Hausdorff | 0.70 [0.40, 1.00] | 0.58 [-0.14, 1.29] | -1.62 |
|  |  |  | AHV | 0.78 [0.46, 1.09] | -0.84 [-1.57, -0.11]* | -2.30 |
|  |  | 48 | ADNS | 0.44 [0.14, 0.75] | 0.11 [-0.44, 0.65] | -0.39 |
|  |  |  | Euclidean | 0.40 [0.15, 0.65] | 0.04 [-0.50, 0.58] | -0.15 |
|  |  |  | 1-D Fréchet | 0.40 [0.15, 0.64] | 0.06 [-0.48, 0.60] | -0.21 |
|  |  |  | N-D Fréchet | 0.63 [0.34, 0.93] | 0.36 [-0.18, 0.91] | -1.32 |
|  |  |  | 1-D Hausdorff | 0.38 [0.10, 0.66] | 0.06 [-0.48, 0.60] | -0.21 |
|  |  |  | N-D Hausdorff | 0.40 [0.16, 0.63] | -0.04 [-0.58, 0.50] | -0.14 |
|  |  |  | AHV | 0.72 [0.46, 0.98] | -0.65 [-1.20, -0.10]* | -2.34 |
|  | MCI | 12 | ADNS | 0.77 [0.65, 0.90] | 1.08 [0.75, 1.42]*** | -6.39 |
|  |  |  | Euclidean | 0.73 [0.55, 0.90] | 1.00 [0.67, 1.33]*** | -5.90 |
|  |  |  | 1-D Fréchet | 0.73 [0.57, 0.89] | 0.97 [0.64, 1.30]*** | -5.73 |
|  |  |  | N-D Fréchet | 0.67 [0.49, 0.84] | 0.69 [0.36, 1.02]*** | -4.13 |
|  |  |  | 1-D Hausdorff | 0.72 [0.55, 0.89] | 0.97 [0.64, 1.30]*** | -5.73 |
|  |  |  | N-D Hausdorff | 0.64 [0.45, 0.83] | 0.45 [0.12, 0.77]** | -2.69 |
|  |  |  | AHV | 0.68 [0.51, 0.85] | -0.69 [-1.02, -0.36]*** | -4.14 |
|  |  | 24 | ADNS | 0.76 [0.64, 0.88] | 1.05 [0.77, 1.34]*** | -7.36 |
|  |  |  | Euclidean | 0.68 [0.53, 0.83] | 0.84 [0.56, 1.12]*** | -5.94 |
|  |  |  | 1-D Fréchet | 0.69 [0.55, 0.84] | 0.82 [0.54, 1.09]*** | -5.78 |
|  |  |  | N-D Fréchet | 0.63 [0.47, 0.78] | 0.65 [0.37, 0.92]*** | -4.61 |
|  |  |  | 1-D Hausdorff | 0.70 [0.57, 0.83] | 0.82 [0.54, 1.09]*** | -5.78 |
|  |  |  | N-D Hausdorff | 0.68 [0.53, 0.84] | 0.66 [0.39, 0.94]*** | -4.73 |
|  |  |  | AHV | 0.73 [0.59, 0.86] | -0.85 [-1.12, -0.57]*** | -5.99 |
|  |  | 36 | ADNS | 0.73 [0.62, 0.84] | 0.87 [0.59, 1.16]*** | -5.99 |
|  |  |  | Euclidean | 0.67 [0.50, 0.83] | 0.68 [0.40, 0.97]*** | -4.74 |
|  |  |  | 1-D Fréchet | 0.69 [0.54, 0.84] | 0.70 [0.41, 0.98]*** | -4.82 |
|  |  |  | N-D Fréchet | 0.63 [0.46, 0.81] | 0.44 [0.16, 0.72]** | -3.06 |
|  |  |  | 1-D Hausdorff | 0.69 [0.54, 0.84] | 0.70 [0.41, 0.98]*** | -4.82 |
|  |  |  | N-D Hausdorff | 0.63 [0.52, 0.73] | 0.49 [0.21, 0.77]*** | -3.41 |
|  |  |  | AHV | 0.73 [0.56, 0.90] | -0.86 [-1.14, -0.57]*** | -5.87 |
|  |  | 48 | ADNS | 0.73 [0.57, 0.89] | 0.78 [0.46, 1.10]*** | -4.83 |
|  |  |  | Euclidean | 0.62 [0.42, 0.82] | 0.54 [0.23, 0.86]*** | -3.41 |
|  |  |  | 1-D Fréchet | 0.59 [0.44, 0.75] | 0.52 [0.21, 0.83]** | -3.25 |

|  |  |  |  |  |  |
| --- | --- | --- | --- | --- | --- |
|  |  | N-D Fréchet | 0.57 [0.41, 0.74] | 0.40 [0.09, 0.71]* | -2.54 |
|  |  | 1-D Hausdorff | 0.63 [0.44, 0.83] | 0.52 [0.21, 0.83]** | -3.25 |
|  |  | N-D Hausdorff | 0.60 [0.41, 0.78] | 0.36 [0.05, 0.68]* | -2.30 |
|  |  | AHV | 0.69 [0.51, 0.88] | -0.76 [-1.08, -0.44]*** | -4.71 |

**Supplementary Table 4.** Significant Neuroreader-Based ROIs by Z-Score Ranking and CN Template Values. The 12 significant regions extracted by performing ANOVA for each of the 22 Neuroreader-based regions in the ROI selection cohort are reported in the above table along with corresponding z-scores. Significance was established based on an alpha=0.05, Bonferroni-corrected. Structures are identified by anatomical labels used by Neuroreader. Mean volumes and standard deviations (SD) of the CN template cohort are included for each respective region.

| Neuroreader ROI Label | Z | CN Template Mean Vol (±SD) ( mm<sup>3</sup> ) |
| --- | --- | --- |
| Left Hippocampus | 9.12 | 0.48 (±0.74) |
| Left Temporal Lobe | 8.43 | 0.45 (±0.68) |
| Left Amygdala | 8.43 | 0.60 (±0.79) |
| Right Amygdala | 8.37 | 0.54 (±0.80) |
| Right Hippocampus | 7.82 | 0.45 (±0.71) |
| Right Temporal Lobe | 7.53 | 0.44 (±0.72) |
| Left Parietal Lobe | 6.74 | 0.37 (±0.83) |
| Right Parietal Lobe | 6.55 | 0.40 (±0.93) |
| Left Frontal Lobe | 5.71 | 0.30 (±0.94) |
| Right Frontal Lobe | 5.48 | 0.26 (±0.98) |
| Left Thalamus | 5.17 | 0.33 (±1.01) |
| Right Thalamus | 4.42 | 0.31 (±1.02) |

**Supplemental Table 5.** Baseline Assessment of Sensitivity to Diagnosis Using Neuroreader-Based ROIs. Assessment of AD-NeuroScore on the basis of metric sensitivity to diagnosis at baseline, tested again using the Neuroreader-based ROIs to generate AD-NeuroScore. AHV, computed using the Neuroreader-based ROIs is included for benchmarking. Significant group comparisons are denoted by * to indicate p<0.05, ** to indicate p<0.01, and *** to indicate p<0.001, Holm-Bonferroni corrected.

| Group Comparison | Metric | AUC [95% CI] | Cohen's *d* [95% CI] | z-score |
| --- | --- | --- | --- | --- |
| CN vs MCI | ADNS | 0.64 [0.57, 0.71] | -0.55 [-0.58, -0.52]*** | -7.31 |
|  | AHV | 0.65 [0.56, 0.74] | 0.55 [0.52, 0.58]*** | -7.27 |
| CN vs AD | ADNS | 0.91 [0.86, 0.97] | -2.11 [-2.29, -1.93]*** | -16.67 |
|  | AHV | 0.89 [0.82, 0.95] | 1.72 [1.55, 1.89]*** | -14.27 |
| MCI vs AD | ADNS | 0.81 [0.74, 0.88] | -1.18 [-1.20, -1.15]*** | -11.35 |
|  | AHV | 0.76 [0.67, 0.85] | 0.92 [0.90, 0.94]*** | -9.04 |

**Supplementary Table 6.**
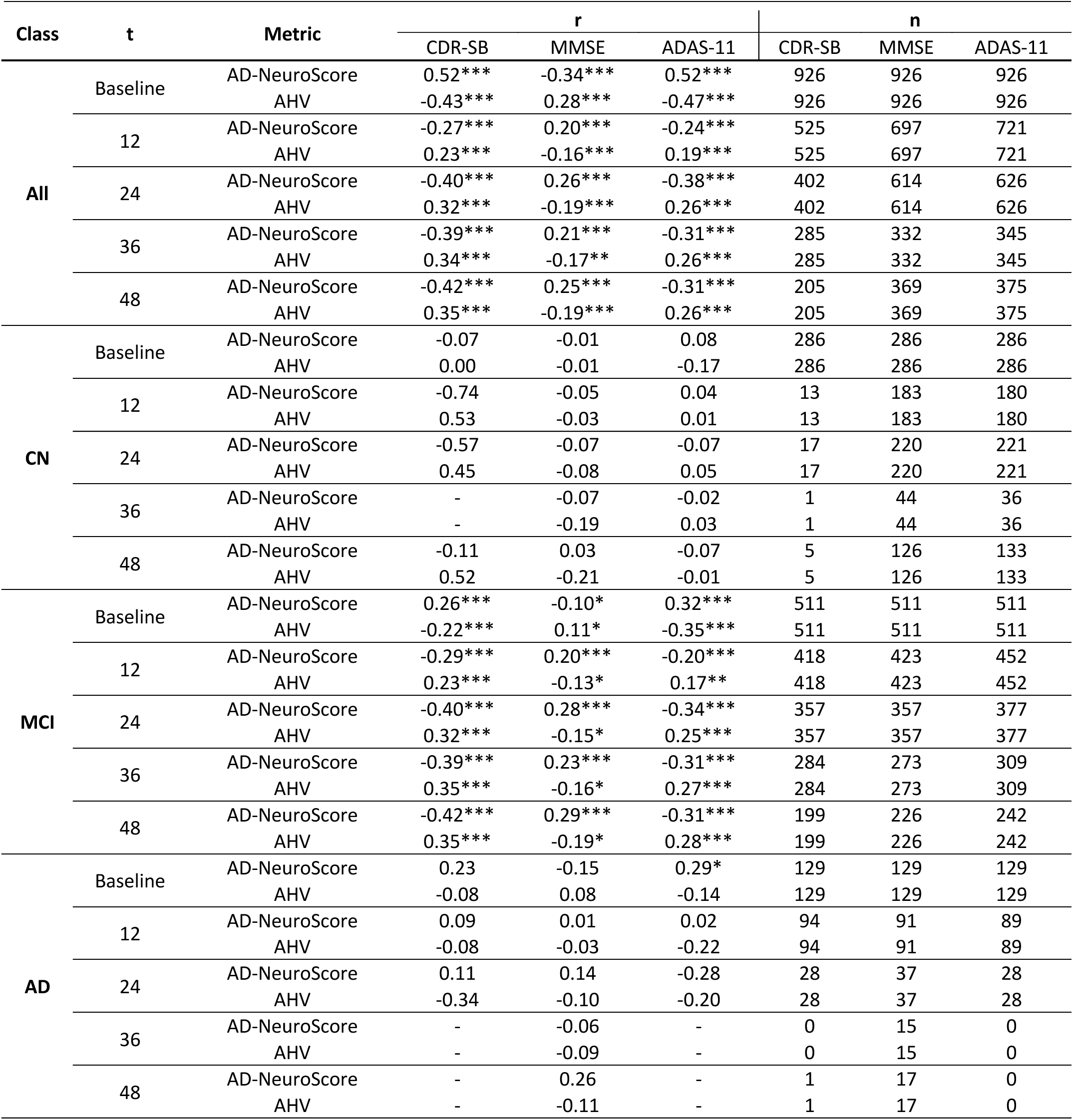
Linear Regression Results using Neuroreader-Based ROIs. Repeated baseline and longitudinal assessment of AD-NeuroScore based on metric association with disease severity (MMSE, ADAS-11, and CDR-SB scores) along with the AHV benchmark, tested with linear regression in the overall experimental cohort and in each diagnostic sub-group, using the Neuroreader-based ROIs to generate AD-NeuroScore and AHV. Significant group comparisons are denoted by * to indicate p<0.05, ** to indicate p<0.01, and *** to indicate p<0.001, Holm-Bonferroni corrected.

**Supplementary Table 7.** Longitudinal Assessment of AD-NeuroScore (ADNS) and AHV based on Diagnosis Transitions Using Neuroreader-Based ROIs. AD-NeuroScore (ADNS) assessment based on ability to predict progression (DTC) at each longitudinal session, using Neuroreader analogous ROIs. Results are stratified by diagnostic group. Two-tailed t-tests were performed for each DTC comparison and z-scores, effect sizes (Cohen’s *d*) with 95% CIs, and AUC-ROC values with 95% confidence intervals were subsequently calculated. Significant group comparisons are denoted by * to indicate p<0.05, ** to indicate p<0.01, and *** to indicate p<0.001, Holm-Bonferroni corrected.

| Comparison |  | t | Metric | AUC [95% CI] | Cohen's *d* [95% CI] | z-score |
| --- | --- | --- | --- | --- | --- | --- |
| Stable vs Decline | All | 12 months | ADNS | 0.66 [0.50, 0.83] | 0.62 [0.33, 0.91]*** | -4.21 |
|  |  |  | AHV | 0.66 [0.53, 0.79] | -0.53 [-0.81, -0.24]** | -3.57 |
|  |  | 24 months | ADNS | 0.72 [0.58, 0.86] | 0.86 [0.62, 1.09]*** | -7.14 |
|  |  |  | AHV | 0.72 [0.62, 0.83] | -0.79 [-1.02, -0.55]*** | -6.58 |
|  |  | 36 months | ADNS | 0.69 [0.58, 0.81] | 0.74 [0.48, 1.00]*** | -5.53 |
|  |  |  | AHV | 0.72 [0.59, 0.84] | -0.79 [-1.05, -0.52]*** | -5.88 |
|  |  | 48 months | ADNS | 0.68 [0.55, 0.82] | 0.77 [0.49, 1.04]*** | -5.53 |
|  |  |  | AHV | 0.71 [0.58, 0.85] | -0.79 [-1.06, -0.51]*** | -5.66 |
|  | CN | 12 months | ADNS | 0.54 [0.14, 0.94] | -0.24 [-0.88, 0.40] | -0.73 |
|  |  |  | AHV | 0.65 [0.27, 1.03] | -0.59 [-1.23, 0.06] | -1.79 |
|  |  | 24 months | ADNS | 0.56 [0.22, 0.89] | 0.36 [-0.10, 0.82] | -1.53 |
|  |  |  | AHV | 0.75 [0.51, 0.99] | -0.79 [-1.26, -0.33]** | -3.34 |
|  |  | 36 months | ADNS | 0.34 [0.06, 0.62] | -0.02 [-0.72, 0.68] | -0.06 |
|  |  |  | AHV | 0.77 [0.48, 1.07] | -0.84 [-1.57, -0.11] | -2.30 |
|  |  | 48 months | ADNS | 0.43 [0.09, 0.77] | 0.11 [-0.44, 0.65] | -0.39 |
|  |  |  | AHV | 0.73 [0.51, 0.94] | -0.65 [-1.20, -0.10] | -2.34 |
|  | MCI | 12 months | ADNS | 0.77 [0.63, 0.91] | 1.07 [0.74, 1.41]*** | -6.33 |
|  |  |  | AHV | 0.69 [0.51, 0.87] | -0.69 [-1.02, -0.36]*** | -4.14 |
|  |  | 24 months | ADNS | 0.77 [0.65, 0.88] | 1.12 [0.84, 1.40]*** | -7.78 |
|  |  |  | AHV | 0.72 [0.59, 0.85] | -0.85 [-1.12, -0.57]*** | -5.99 |
|  |  | 36 months | ADNS | 0.73 [0.61, 0.86] | 0.94 [0.65, 1.23]*** | -6.41 |
|  |  |  | AHV | 0.74 [0.61, 0.86] | -0.86 [-1.14, -0.57]*** | -5.87 |
|  |  | 48 months | ADNS | 0.73 [0.57, 0.89] | 0.82 [0.51, 1.14]*** | -5.09 |
|  |  |  | AHV | 0.69 [0.53, 0.86] | -0.76 p-1.08, -0.44]*** | -4.71 |

## References

Achterberg, H.C., van der Lijn, F., den Heijer, T., Vernooij, M.W., Ikram, M.A., Niessen, W.J., de Bruijne, M., 2014. Hippocampal shape is predictive for the development of dementia in a normal, elderly population. Hum Brain Mapp 35, 2359–2371. https://doi.org/10.1002/HBM.22333

Ahdidan, J., Raji, C.A., DeYoe, E.A., Mathis, J., Noe, K.O., Rimestad, J., Kjeldsen, T.K., Mosegaard, J., Becker, J.T., Lopez, O., 2016. Quantitative Neuroimaging Software for Clinical Assessment of Hippocampal Volumes on MR Imaging. J Alzheimers Dis 49, 723– 732. https://doi.org/10.3233/JAD-150559

Archetti, D., Young, A.L., Oxtoby, N.P., Ferreira, D., Mårtensson, G., Westman, E., Alexander, D.C., Frisoni, G.B., Redolfi, A., 2021. Inter-cohort validation of sustain model for alzheimer’s disease. Front Big Data 4, 30. https://doi.org/10.3389/FDATA.2021.661110/BIBTEX

Attier-Zmudka, J., Sérot, J.M., Valluy, J., Saffarini, M., Macaret, A.S., Diouf, M., Dao, S., Douadi, Y., Piotr Malinowski, K., Balédent, O., 2019. Decreased Cerebrospinal Fluid Flow Is Associated With Cognitive Deficit in Elderly Patients. Front Aging Neurosci 11. https://doi.org/10.3389/FNAGI.2019.00087

Balsis, S., Benge, J.F., Lowe, D.A., Geraci, L., Doody, R.S., 2015. How Do Scores on the ADAS-Cog, MMSE, and CDR-SOB Correspond? Clin Neuropsychol 29, 1002–1009. https://doi.org/10.1080/13854046.2015.1119312

Braak, H., Braak, Eva, Braak, (H, Braak, E, 1997. Staging of Alzheimer-Related Cortical Destruction. Int Psychogeriatr 9, 257–261. https://doi.org/10.1017/S1041610297004973

Brewer, J.B., Magda, S., Airriess, C., Smith, M.E., 2009. Fully-Automated Quantification of Regional Brain Volumes for Improved Detection of Focal Atrophy in Alzheimer Disease. American Journal of Neuroradiology 30, 578–580. https://doi.org/10.3174/AJNR.A1402

Cabeza, R., Ciaramelli, E., Olson, I.R., Moscovitch, M., 2008. Parietal Cortex and Episodic Memory: An Attentional Account. Nat Rev Neurosci 9, 613. https://doi.org/10.1038/NRN2459

Casanova, R., Barnard, R.T., Gaussoin, S.A., Saldana, S., Hayden, K.M., Manson, J.A.E., Wallace, R.B., Rapp, S.R., Resnick, S.M., Espeland, M.A., Chen, J.C., 2018a. Using high-dimensional machine learning methods to estimate an anatomical risk factor for Alzheimer’s disease across imaging databases. Neuroimage 183, 401–411. https://doi.org/10.1016/J.NEUROIMAGE.2018.08.040

Casanova, R., Hsu, F.C., Sink, K.M., Rapp, S.R., Williamson, J.D., Resnick, S.M., Espeland, M.A., 2013. Alzheimer’s Disease Risk Assessment Using Large-Scale Machine Learning Methods. PLoS One 8, e77949. https://doi.org/10.1371/JOURNAL.PONE.0077949

Cavedo, E., Tran, P., Thoprakarn, U., Martini, J.B., Movschin, A., Delmaire, C., Gariel, F., Heidelberg, D., Pyatigorskaya, N., Ströer, S., Krolak-Salmon, P., Cotton, F., dos Santos, C.L., Dormont, D., 2022. Validation of an automatic tool for the rapid measurement of brain atrophy and white matter hyperintensity: QyScore®. Eur Radiol 32, 2949–2961. https://doi.org/10.1007/S00330-021-08385-9

Chow, N., Hwang, K.S., Hurtz, S., Green, A.E., Somme, J.H., Thompson, P.M., Elashoff, D.A., Jack, C.R., Weiner, M., Apostolova, L.G., 2015. Comparing 3T and 1.5T MRI for Mapping Hippocampal Atrophy in the Alzheimer’s Disease Neuroimaging Initiative. American Journal of Neuroradiology 36, 653–660. https://doi.org/10.3174/AJNR.A4228

Coupé, P., Fonov, V.S., Bernard, C., Zandifar, A., Eskildsen, S.F., Helmer, C., Manjón, J. v., Amieva, H., Dartigues, J.F., Allard, M., Catheline, G., Collins, D.L., 2015a. Detection of Alzheimer’s disease signature in MR images seven years before conversion to dementia: Toward an early individual prognosis. Hum Brain Mapp 36, 4758–4770. https://doi.org/10.1002/HBM.22926

Csernansky, J.G., Wang, L., Joshi, S.C., Tilak Ratnanather, J., Miller, M.I., 2004. Computational anatomy and neuropsychiatric disease: probabilistic assessment of variation and statistical inference of group difference, hemispheric asymmetry, and time-dependent change. Neuroimage 23, S56–S68. https://doi.org/10.1016/J.NEUROIMAGE.2004.07.025

Dale, A., Fischl, B., Sereno, M.I., 1999. Cortical Surface-Based Analysis: I. Segmentation and Surface Reconstruction. Neuroimage 9, 179–194.

Davatzikos, C., Xu, F., An, Y., Fan, Y., Resnick, S.M., 2009. Longitudinal progression of Alzheimer’s-like patterns of atrophy in normal older adults: the SPARE-AD index. Brain 132, 2026–2035. https://doi.org/10.1093/BRAIN/AWP091

Desikan, R.S., Ségonne, F., Fischl, B., Quinn, B.T., Dickerson, B.C., Blacker, D., Buckner, R.L., Dale, A.M., Maguire, R.P., Hyman, B.T., Albert, M.S., Killiany, R.J., 2006. An automated labeling system for subdividing the human cerebral cortex on MRI scans into gyral based regions of interest. Neuroimage 31, 968–980. https://doi.org/DOI: 10.1016/j.neuroimage.2006.01.021

Diciotti, S., Ginestroni, A., Bessi, V., Giannelli, M., Tessa, C., Bracco, L., Mascalchi, M., Toschi, N., 2012a. Identification of mild Alzheimer’s disease through automated classification of structural MRI features. Annu Int Conf IEEE Eng Med Biol Soc 2012, 428–431. https://doi.org/10.1109/EMBC.2012.6345959

Dickerson, B.C., Goncharova, I., Sullivan, M.P., Forchetti, C., Wilson, R.S., Bennett, D.A., Beckett, L.A., DeToledo-Morrell, L., 2001. MRI-derived entorhinal and hippocampal atrophy in incipient and very mild Alzheimer’s disease. Neurobiol Aging 22, 747–754. https://doi.org/10.1016/S0197-4580(01)00271-8

Dukart, J., Mueller, K., Barthel, H., Villringer, A., Sabri, O., Schroeter, M.L., 2013. Meta-analysis based SVM classification enables accurate detection of Alzheimer’s disease across different clinical centers using FDG-PET and MRI. Psychiatry Res 212, 230–236. https://doi.org/10.1016/J.PSCYCHRESNS.2012.04.007

Dumitrescu, A., Rote, G., 2004. On the Fréchet distance of a set of curves.

Elahi, S., Bachman, A.H., Lee, S.H., Sidtis, J.J., Ardekani, B.A., 2015. Corpus Callosum Atrophy Rate in Mild Cognitive Impairment and Prodromal Alzheimer’s Disease. J Alzheimers Dis 45, 921. https://doi.org/10.3233/JAD-142631

Ezzati, A., Zammit, A.R., Harvey, D.J., Habeck, C., Hall, C.B., Lipton, R.B., 2019. Optimizing Machine Learning Methods to Improve Predictive Models of Alzheimer’s Disease. J Alzheimers Dis 71, 1027. https://doi.org/10.3233/JAD-190262

Fischl, B., Liu, A., Dale, A. ∼M., 2001. Automated manifold surgery: constructing geometrically accurate and topologically correct models of the human cerebral cortex. IEEE Medical Imaging 20, 70–80.

Fischl, B., Salat, D. ∼H., Busa, E., Albert, M., Dieterich, M., Haselgrove, C., van der Kouwe, A., Killiany, R., Kennedy, D., Klaveness, S., Montillo, A., Makris, N., Rosen, B., Dale, A. ∼M., 2002. Whole brain segmentation: automated labeling of neuroanatomical structures in the human brain. Neuron 33, 341–355.

Fischl, B., Salat, D.H., van der Kouwe, A.J.W., Makris, N., Ségonne, F., Quinn, B.T., Dale, A.M., 2004a. Sequence-independent segmentation of magnetic resonance images. Neuroimage 23, S69–S84. https://doi.org/DOI: 10.1016/j.neuroimage.2004.07.016

Fischl, B., Sereno, M.I., Dale, A., 1999a. Cortical Surface-Based Analysis: II: Inflation, Flattening, and a Surface-Based Coordinate System. Neuroimage 9, 195–207.

Fischl, B., Sereno, M.I., Tootell, R.B.H., Dale, A.M., 1999b. High-resolution intersubject averaging and a coordinate system for the cortical surface. Hum Brain Mapp 8, 272–284. https://doi.org/10.1002/(SICI)1097-0193(1999)8:4<272::AID-HBM10>3.0.CO;2-4

Fischl, B., van der Kouwe, A., Destrieux, C., Halgren, E., Ségonne, F., Salat, D.H., Busa, E., Seidman, L.J., Goldstein, J., Kennedy, D., Caviness, V., Makris, N., Rosen, B., Dale, A.M., 2004b. Automatically Parcellating the Human Cerebral Cortex. Cerebral Cortex 14, 11–22. https://doi.org/10.1093/cercor/bhg087

Folstein, M.F., Folstein, S.E., McHugh, P.R., 1975. “Mini-mental state”. A practical method for grading the cognitive state of patients for the clinician. J Psychiatr Res 12, 189–198. https://doi.org/10.1016/0022-3956(75)90026-6

Fonteijn, H.M., Modat, M., Clarkson, M.J., Barnes, J., Lehmann, M., Hobbs, N.Z., Scahill, R.I., Tabrizi, S.J., Ourselin, S., Fox, N.C., Alexander, D.C., 2012. An event-based model for disease progression and its application in familial Alzheimer’s disease and Huntington’s disease. Neuroimage 60, 1880–1889. https://doi.org/10.1016/J.NEUROIMAGE.2012.01.062

Frisoni, G.B., Laakso, M.P., Beltramello, A., Geroldi, C., Bianchetti, A., Soininen, H., Trabucchi, M., 1999. Hippocampal and entorhinal cortex atrophy in frontotemporal dementia and Alzheimer’s disease. Neurology 52, 91–100. https://doi.org/10.1212/WNL.52.1.91

Giesel, F.L., Hahn, H.K., Thomann, P.A., Widjaja, E., Wignall, E., von Tengg-Kobligk, H., Pantel, J., Griffiths, P.D., Peitgen, H.O., Schroder, J., Essig, M., n.d. Temporal Horn Index and Volume of Medial Temporal Lobe Atrophy Using a New Semiautomated Method for Rapid and Precise Assessment.

Gómez-Isla, T., Price, J.L., McKeel, D.W., Morris, J.C., Growdon, J.H., Hyman, B.T., 1996a. Profound loss of layer II entorhinal cortex neurons occurs in very mild Alzheimer’s disease. J Neurosci 16, 4491–4500. https://doi.org/10.1523/JNEUROSCI.16-14-04491.1996

Gosche, K.M., Mortimer, J.A., Smith, C.D., Markesbery, W.R., Snowdon, D.A., 2002. Hippocampal volume as an index of Alzheimer neuropathology. Neurology 58, 1476–1482. https://doi.org/10.1212/WNL.58.10.1476

Grochowalski, J.H., Liu, Y., Siedlecki, K.L., 2015. Aging, Neuropsychology, and Cognition A Journal on Normal and Dysfunctional Development Examining the reliability of ADAS-Cog change scores. https://doi.org/10.1080/13825585.2015.1127320

Gutman, B.A., Hua, X., Rajagopalan, P., Chou, Y.Y., Wang, Y., Yanovsky, I., Toga, A.W., Jack, C.R., Weiner, M.W., Thompson, P.M., 2013. Maximizing power to track Alzheimer’s disease and MCI progression by LDA-based weighting of longitudinal ventricular surface features. Neuroimage 70, 386. https://doi.org/10.1016/J.NEUROIMAGE.2012.12.052

Harper, L., Bouwman, F., Burton, E.J., Barkhof, F., Scheltens, P., O’Brien, J.T., Fox, N.C., Ridgway, G.R., Schott, J.M., 2017. Patterns of atrophy in pathologically confirmed dementias: a voxelwise analysis. J Neurol Neurosurg Psychiatry 88, 908–916. https://doi.org/10.1136/jnnp-2016-314978

Harris, C.R., Millman, K.J., van der Walt, S.J., Gommers, R., Virtanen, P., Cournapeau, D., Wieser, E., Taylor, J., Berg, S., Smith, N.J., Kern, R., Picus, M., Hoyer, S., van Kerkwijk, M.H., Brett, M., Haldane, A., del Río, J.F., Wiebe, M., Peterson, P., Gérard-Marchant, P., Sheppard, K., Reddy, T., Weckesser, W., Abbasi, H., Gohlke, C., Oliphant, T.E., 2020. Array programming with NumPy. Nature 585, 357–362. https://doi.org/10.1038/S41586-020-2649-2

Hotelling, H., 1947. Multivariate Quality Control Illustrated by Air Testing of Sample Bombsights., in: Eisenhart, C., Hastay, M.W., Wallis, W.A. (Eds.), Techniques of Statistical Analysis. McGraw Hill, New York, pp. 111–184.

Hua, X., Hibar, D.P., Ching, C.R.K., Boyle, C.P., Rajagopalan, P., Gutman, B.A., Leow, A.D., Toga, A.W., Jack, C.R., Harvey, D., Weiner, M.W., Thompson, P.M., 2013. Unbiased tensor-based morphometry: Improved robustness and sample size estimates for Alzheimer’s disease clinical trials. Neuroimage 66, 648. https://doi.org/10.1016/J.NEUROIMAGE.2012.10.086

Hughes, C.P., Berg, L., Danziger, W.L., Coben, L.A., Martin, R.L., 1982. A new clinical scale for the staging of dementia. Br J Psychiatry 140, 566–572. https://doi.org/10.1192/BJP.140.6.566

Hwang, E.J., Park, C.M., 2020. Clinical Implementation of Deep Learning in Thoracic Radiology: Potential Applications and Challenges. Korean J Radiol 21, 511. https://doi.org/10.3348/KJR.2019.0821

Jack, C.R., Bennett, D.A., Blennow, K., Carrillo, M.C., Dunn, B., Haeberlein, S.B., Holtzman, D.M., Jagust, W., Jessen, F., Karlawish, J., Liu, E., Molinuevo, J.L., Montine, T., Phelps, C., Rankin, K.P., Rowe, C.C., Scheltens, P., Siemers, E., Snyder, H.M., Sperling, R., Elliott, C., Masliah, E., Ryan, L., Silverberg, N., 2018. NIA-AA Research Framework: Toward a biological definition of Alzheimer’s disease. Alzheimers Dement 14, 535–562. https://doi.org/10.1016/J.JALZ.2018.02.018

Jack, C.R., Bernstein, M.A., Fox, N.C., Thompson, P., Alexander, G., Harvey, D., Borowski, B., Britson, P.J., Whitwell, J.L., Ward, C., Dale, A.M., Felmlee, J.P., Gunter, J.L., Hill, D.L.G., Killiany, R., Schuff, N., Fox-Bosetti, S., Lin, C., Studholme, C., DeCarli, C.S., Krueger, G., Ward, H.A., Metzger, G.J., Scott, K.T., Mallozzi, R., Blezek, D., Levy, J., Debbins, J.P., Fleisher, A.S., Albert, M., Green, R., Bartzokis, G., Glover, G., Mugler, J., Weiner, M.W., 2008. The Alzheimer’s Disease Neuroimaging Initiative (ADNI): MRI methods. J Magn Reson Imaging 27, 685–691. https://doi.org/10.1002/JMRI.21049

Jack, C.R., Dickson, D.W., Parisi, J.E., Xu, Y.C., Cha, R.H., O’Brien, P.C., Edland, S.D., Smith, G.E., Boeve, B.F., Tangalos, E.G., Kokmen, E., Petersen, R.C., 2002. Antemortem MRI Findings Correlate with Hippocampal Neuropathology in Typical Aging and Dementia. Neurology 58, 750. https://doi.org/10.1212/WNL.58.5.750

Jack, C.R., Petersen, R.C., Xu, Y.C., Waring, S.C., O’Brien, P.C., Tangalos, E.G., Smith, G.E., Ivnik, R.J., Kokmen, E., 1997. Medial temporal atrophy on MRI in normal aging and very mild Alzheimer’s disease. Neurology 49, 786–794. https://doi.org/10.1212/WNL.49.3.786

Jacobs, H.I.L., van Boxtel, M.P.J., Jolles, J., Verhey, F.R.J., Uylings, H.B.M., 2012a. Parietal cortex matters in Alzheimer’s disease: An overview of structural, functional and metabolic findings. Neurosci Biobehav Rev 36, 297–309. https://doi.org/10.1016/J.NEUBIOREV.2011.06.009

Jacobs, H.I.L., van Boxtel, M.P.J., Jolles, J., Verhey, F.R.J., Uylings, H.B.M., 2012b. Parietal cortex matters in Alzheimer’s disease: an overview of structural, functional and metabolic findings. Neurosci Biobehav Rev 36, 297–309. https://doi.org/10.1016/J.NEUBIOREV.2011.06.009

Jovicich, J., Czanner, S., Greve, D., Haley, E., van der Kouwe, A., Gollub, R., Kennedy, D., Schmitt, F., Brown, G., MacFall, J., Fischl, B., Dale, A., 2006. Reliability in multi-site structural MRI studies: Effects of gradient non-linearity correction on phantom and human data. Neuroimage 30, 436–443. https://doi.org/DOI: 10.1016/j.neuroimage.2005.09.046

Khan, W., Westman, E., Jones, N., Wahlund, L.O., Mecocci, P., Vellas, B., Tsolaki, M., Kłoszewska, I., Soininen, H., Spenger, C., Lovestone, S., Muehlboeck, J.S., Simmons, A., 2015. Automated Hippocampal Subfield Measures as Predictors of Conversion from Mild Cognitive Impairment to Alzheimer’s Disease in Two Independent Cohorts. Brain Topogr 28, 746–759. https://doi.org/10.1007/S10548-014-0415-1

Kochunov, P., Ma, Y., Hatch, K.S., Schmaal, L., Jahanshad, N., Thompson, P.M., Adhikari, B.M., Bruce, H., Chiappelli, J., van der Vaart, A., Goldwaser, E.L., Sotiras, A., Ma, T., Chen, S., Nichols, T.E., Hong, L.E., 2022. Separating Clinical and Subclinical Depression by Big Data Informed Structural Vulnerability Index and Its impact on Cognition: ENIGMA Dot Product. Pac Symp Biocomput 27, 133. https://doi.org/10.1142/9789811250477_0013

Kochunov, P., Ryan, M.C., Yang, Q., Hatch, K.S., Zhu, A., Thomopoulos, S.I., Jahanshad, N., Schmaal, L., Thompson, P.M., Chen, S., Du, X., Adhikari, B.M., Bruce, H., Hare, S., Goldwaser, E.L., Kvarta, M.D., Nichols, T.E., Hong, L.E., 2021. Comparison of regional brain deficit patterns in common psychiatric and neurological disorders as revealed by big data. Neuroimage Clin 29, 102574. https://doi.org/10.1016/J.NICL.2021.102574

Koikkalainen, J., Rhodius-Meester, H., Tolonen, A., Barkhof, F., Tijms, B., Lemstra, A.W., Tong, T., Guerrero, R., Schuh, A., Ledig, C., Rueckert, D., Soininen, H., Remes, A.M., Waldemar, G., Hasselbalch, S., Mecocci, P., van der Flier, W., Lötjönen, J., 2016. Differential diagnosis of neurodegenerative diseases using structural MRI data. Neuroimage Clin 11, 435–449. https://doi.org/10.1016/J.NICL.2016.02.019

Laakso, M.P., Partanen, K., Riekkinen, P., Lehtovirta, M., Helkala, E.L., Hallikainen, M., Hänninen, T., Vainio, P., Soininen, H., 1996. Hippocampal volumes in Alzheimer’s disease, Parkinson’s disease with and without dementia, and in vascular dementia: An MRI study. Neurology 46, 678–681. https://doi.org/10.1212/WNL.46.3.678

Laakso, M.P., Soininen, H., Partanen, K., Lehtovirta, M., Hallikainen, M., Hänninen, T., Helkala, E.L., Vainio, P., Riekkinen, P.J., 1998. MRI of the hippocampus in Alzheimer’s disease: sensitivity, specificity, and analysis of the incorrectly classified subjects. Neurobiol Aging 19, 23–31. https://doi.org/10.1016/S0197-4580(98)00006-2

Lindeboom, J., Weinstein, H., 2004. Neuropsychology of cognitive ageing, minimal cognitive impairment, Alzheimer’s disease, and vascular cognitive impairment. Eur J Pharmacol 490, 83–86. https://doi.org/10.1016/J.EJPHAR.2004.02.046

Lu, B., Li, H.-X., Chang, Z.-K., Li, L., Chen, N.-X., Zhu, Z.-C., Zhou, H.-X., Li, X.-Y., Wang, Y.-W., Cui, S.-X., Deng, Z.-Y., Fan, Z., Yang, H., Chen, X., Thompson, P.M., Castellanos, F.X., Yan, C.-G., Initiative, for the A.D.N., 2021. A Practical Alzheimer Disease Classifier via Brain Imaging-Based Deep Learning on 85,721 Samples. bioRxiv 2020.08.18.256594. https://doi.org/10.1101/2020.08.18.256594

Ma, D., Popuri, K., Bhalla, M., Sangha, O., Lu, D., Cao, J., Jacova, C., Wang, L., Beg, M.F., 2019. Quantitative assessment of field strength, total intracranial volume, sex, and age effects on the goodness of harmonization for volumetric analysis on the ADNI database. Hum Brain Mapp 40, 1507–1527. https://doi.org/10.1002/HBM.24463

Maiseli, B.J., 2021. Hausdorff Distance with Outliers and Noise Resilience Capabilities. SN Comput Sci 2. https://doi.org/10.1007/S42979-021-00737-Y

Mazziotta, J.C., Toga, A.W., Evans, A., Fox, P., Lancaster, J., 1995a. A Probabilistic Atlas of the Human Brain: Theory and Rationale for Its Development: The International Consortium for Brain Mapping (ICBM). Neuroimage 2, 89–101. https://doi.org/10.1006/NIMG.1995.1012

Mazziotta, J.C., Toga, A.W., Evans, A.C., Fox, P.T., Lancaster, J.L., 1995b. Digital brain atlases. Trends Neurosci 18, 210–211. https://doi.org/10.1016/0166-2236(95)93904-C

Morris, J.C., 1993. The Clinical Dementia Rating (CDR). Neurology 43, 2412–2412-a. https://doi.org/10.1212/WNL.43.11.2412-A

Mueller, S.G., Weiner, M.W., Thal, L.J., Petersen, R.C., Jack, C., Jagust, W., Trojanowski, J.Q., Toga, A.W., Beckett, L., 2005a. The Alzheimer’s disease neuroimaging initiative. Neuroimaging Clin N Am 15, 869–877. https://doi.org/10.1016/J.NIC.2005.09.008

Mueller, S.G., Weiner, M.W., Thal, L.J., Petersen, R.C., Jack, C.R., Jagust, W., Trojanowski, J.Q., Toga, A.W., Beckett, L., 2005b. Ways toward an early diagnosis in Alzheimer’s disease: the Alzheimer’s Disease Neuroimaging Initiative (ADNI). Alzheimers Dement 1, 55–66. https://doi.org/10.1016/J.JALZ.2005.06.003

Mukherji, D., Mukherji, M., Mukherji, N., Initiative, A.D.N., 2021. Early Detection of Alzheimer’s Disease with Low-Cost Neuropsychological Tests: A Novel Predict-Diagnose Approach using Recurrent Neural Networks. medRxiv 2021.01.17.21249822. https://doi.org/10.1101/2021.01.17.21249822

Nestor, Sean M., Rupsingh, R., Borrie, M., Smith, M., Accomazzi, V., Wells, J.L., Fogarty, J., Bartha, R., 2008. Ventricular enlargement as a possible measure of Alzheimer’s disease progression validated using the Alzheimer’s disease neuroimaging initiative database. Brain 131, 2443–2454. https://doi.org/10.1093/BRAIN/AWN146

Nestor, Sean M, Rupsingh, R., Borrie, M., Smith, M., Accomazzi, V., Wells, J.L., Fogarty, J., Bartha, R., Initiative, the A.D.N., 2008. Ventricular enlargement as a possible measure of Alzheimer’s disease progression validated using the Alzheimer’s disease neuroimaging initiative database. Brain 131, 2443–2454. https://doi.org/10.1093/BRAIN/AWN146

Nie, X., Sun, Y., Wan, S., Zhao, H., Liu, R., Li, X., Wu, S., Nedelska, Z., Hort, J., Qing, Z., Xu, Y., Zhang, B., 2017. Subregional Structural Alterations in Hippocampus and Nucleus Accumbens Correlate with the Clinical Impairment in Patients with Alzheimer’s Disease Clinical Spectrum: Parallel Combining Volume and Vertex-Based Approach. Front Neurol 8. https://doi.org/10.3389/FNEUR.2017.00399

O’Bryant, S.E., Waring, S.C., Cullum, C.M., Hall, J., Lacritz, L., Massman, P.J., Lupo, P.J., Reisch, J.S., Doody, R., 2008. Staging Dementia Using Clinical Dementia Rating Scale Sum of Boxes Scores: A Texas Alzheimer’s Research Consortium Study. Arch Neurol 65, 1091. https://doi.org/10.1001/ARCHNEUR.65.8.1091

Ott, B.R., Cohen, R.A., Gongvatana, A., Okonkwo, O.C., Johanson, C.E., Stopa, E.G., Donahue, J.E., Silverberg, G.D., 2010. Brain ventricular volume and cerebrospinal fluid biomarkers of Alzheimer’s disease. J Alzheimers Dis 20, 647. https://doi.org/10.3233/JAD-2010-1406

Petersen, R.C., Aisen, P.S., Beckett, L.A., Donohue, M.C., Gamst, A.C., Harvey, D.J., Jack, C.R., Jagust, W.J., Shaw, L.M., Toga, A.W., Trojanowski, J.Q., Weiner, M.W., 2010. Alzheimer’s Disease Neuroimaging Initiative (ADNI): Clinical characterization. Neurology 74, 201. https://doi.org/10.1212/WNL.0B013E3181CB3E25

Pinto, M.F., Leal, A., Lopes, F., Pais, J., Dourado, A., Sales, F., Martins, P., Teixeira, C.A., 2022. On the clinical acceptance of black-box systems for EEG seizure prediction. Epilepsia Open 7. https://doi.org/10.1002/EPI4.12597

Popuri, K., Ma, D., Wang, L., Beg, M.F., 2020a. Using machine learning to quantify structural MRI neurodegeneration patterns of Alzheimer’s disease into dementia score: Independent validation on 8,834 images from ADNI, AIBL, OASIS, and MIRIAD databases. Hum Brain Mapp 41, 4127–4147. https://doi.org/10.1002/HBM.25115

Rabinovici, G.D., Seeley, W.W., Kim, E.J., Gorno-Tempini, M.L., Rascovsky, K., Pagliaro, T.A., Allison, S.C., Halabi, C., Kramer, J.H., Johnson, J.K., Weiner, M.W., Forman, M.S., Trojanowski, J.Q., Dearmond, S.J., Miller, B.L., Rosen, H.J., 2007. Distinct MRI Atrophy Patterns in Autopsy-Proven Alzheimer’s Disease and Frontotemporal Lobar Degeneration. Am J Alzheimers Dis Other Demen 22, 474. https://doi.org/10.1177/1533317507308779

Rallabandi, V. P.Subramanyam, Tulpule, K., Gattu, M., 2020. Automatic classification of cognitively normal, mild cognitive impairment and Alzheimer’s disease using structural MRI analysis. Inform Med Unlocked 18, 100305. https://doi.org/10.1016/J.IMU.2020.100305

Reuter, M., Rosas, H.D., Fischl, B., 2010. Highly Accurate Inverse Consistent Registration: A Robust Approach. Neuroimage 53, 1181–1196. https://doi.org/10.1016/j.neuroimage.2010.07.020

Rosen, W.G., Mohs, R.C., Davis, K.L., 1984. A new rating scale for Alzheimer’s disease. Am J Psychiatry 141. https://doi.org/10.1176/AJP.141.11.1356

Salvatore, C., Cerasa, A., Castiglioni, I., 2018. MRI characterizes the progressive course of AD and predicts conversion to Alzheimer’s dementia 24 months before probable diagnosis. Front Aging Neurosci 10, 135. https://doi.org/10.3389/FNAGI.2018.00135/BIBTEX

Schott, J.M., Price, S.L., Frost, C., Whitwell, J.L., Rossor, M.N., Fox, N.C., 2005. Measuring atrophy in Alzheimer disease. Neurology 65, 119–124. https://doi.org/10.1212/01.WNL.0000167542.89697.0F

Segonne, F., Dale, A.M., Busa, E., Glessner, M., Salat, D., Hahn, H.K., Fischl, B., 2004. A hybrid approach to the skull stripping problem in MRI. Neuroimage 22, 1060–1075. https://doi.org/DOI: 10.1016/j.neuroimage.2004.03.032

Segonne, F., Pacheco, J., Fischl, B., 2007. Geometrically accurate topology-correction of cortical surfaces using nonseparating loops. IEEE Trans Med Imaging 26, 518–529.

Silbert, L.C., Quinn, J.F., Moore, M.M., Corbridge, E., Ball, M.J., Murdoch, G., Sexton, G., Kaye, J.A., 2003. Changes in premorbid brain volume predict Alzheimer’s disease pathology. Neurology 61, 487–492. https://doi.org/10.1212/01.WNL.0000079053.77227.14

Sørensen, L., Igel, C., Liv Hansen, N., Osler, M., Lauritzen, M., Rostrup, E., Nielsen, M., 2016. Early detection of Alzheimer’s disease using MRI hippocampal texture. Hum Brain Mapp 37, 1148–1161. https://doi.org/10.1002/HBM.23091

Stein, J.L., Medland, S.E., Vasquez, A.A., Hibar, D.P., Senstad, R.E., Winkler, A.M., Toro, R., Appel, K., Bartecek, R., Bergmann, Ø., Bernard, M., Brown, A.A., Cannon, D.M., Chakravarty, M.M., Christoforou, A., Domin, M., Grimm, O., Hollinshead, M., Holmes, A.J., Homuth, G., Hottenga, J.-J., Langan, C., Lopez, L.M., Hansell, N.K., Hwang, K.S., Kim, S., Laje, G., Lee, P.H., Liu, X., Loth, E., Lourdusamy, A., Mattingsdal, M., Mohnke, S., Maniega, S.M., Nho, K., Nugent, A.C., O’Brien, C., Papmeyer, M., Pütz, B., Ramasamy, A., Rasmussen, J., Rijpkema, M., Risacher, S.L., Roddey, J.C., Rose, E.J., Ryten, M., Shen, L., Sprooten, E., Strengman, E., Teumer, A., Trabzuni, D., Turner, J., van Eijk, K., van Erp, T.G.M., van Tol, M.-J., Wittfeld, K., Wolf, C., Woudstra, S., Aleman, A., Alhusaini, S., Almasy, L., Binder, E.B., Brohawn, D.G., Cantor, R.M., Carless, M.A., Corvin, A., Czisch, M., Curran, J.E., Davies, G., de Almeida, M.A.A., Delanty, N., Depondt, C., Duggirala, R., Dyer, T.D., Erk, S., Fagerness, J., Fox, P.T., Freimer, N.B., Gill, M., Göring, H.H.H., Hagler, D.J., Hoehn, D., Holsboer, F., Hoogman, M., Hosten, N., Jahanshad, N., Johnson, M.P., Kasperaviciute, D., Kent Jr, J.W., Kochunov, P., Lancaster, J.L., Lawrie, S.M., Liewald, D.C., Mandl, R., Matarin, M., Mattheisen, M., Meisenzahl, E., Melle, I., Moses, E.K., Mühleisen, T.W., Nauck, M., Nöthen, M.M., Olvera, R.L., Pandolfo, M., Pike, G.B., Puls, R., Reinvang, I., Rentería, M.E., Rietschel, M., Roffman, J.L., Royle, N.A., Rujescu, D., Savitz, J., Schnack, H.G., Schnell, K., Seiferth, N., Smith, C., Steen, V.M., Valdés Hernández, M.C., van den Heuvel, M., van der Wee, N.J., van Haren, N.E.M., Veltman, J.A., Völzke, H., Walker, R., Westlye, L.T., Whelan, C.D., Agartz, I., Boomsma, D.I., Cavalleri, G.L., Dale, A.M., Djurovic, S., Drevets, W.C., Hagoort, P., Hall, J., Heinz, A., Jack Jr, C.R., Foroud, T.M., le Hellard, S., Macciardi, F., Montgomery, G.W., Poline, J.B., Porteous, D.J., Sisodiya, S.M., Starr, J.M., Sussmann, J., Toga, A.W., Veltman, D.J., Walter, H., Weiner, M.W., Initiative, A.D.N., Consortium, E., Consortium, I., Group, S.Y.S., Bis, J.C., Ikram, M.A., Smith, A. v, Gudnason, V., Tzourio, C., Vernooij, M.W., Launer, L.J., DeCarli, C., Seshadri, S., Consortium, C. for H. and A.R. in G.E., Andreassen, O.A., Apostolova, L.G., Bastin, M.E., Blangero, J., Brunner, H.G., Buckner, R.L., Cichon, S., Coppola, G., de Zubicaray, G.I., Deary, I.J., Donohoe, G., de Geus, E.J.C., Espeseth, T., Fernández, G., Glahn, D.C., Grabe, H.J., Hardy, J., Hulshoff Pol, H.E., Jenkinson, M., Kahn, R.S., McDonald, C., McIntosh, A.M., McMahon, F.J., McMahon, K.L., Meyer-Lindenberg, A., Morris, D.W., Müller-Myhsok, B., Nichols, T.E., Ophoff, R.A., Paus, T., Pausova, Z., Penninx, B.W., Potkin, S.G., Sämann, P.G., Saykin, A.J., Schumann, G., Smoller, J.W., Wardlaw, J.M., Weale, M.E., Martin, N.G., Franke, B., Wright, M.J., Thompson, P.M., Consortium, E.N.I.G. through M.-A., 2012. Identification of common variants associated with human hippocampal and intracranial volumes. Nat Genet 44, 552–561. https://doi.org/10.1038/ng.2250

Thompson, P.M., Hayashi, K.M., de Zubicaray, G., Janke, A.L., Rose, S.E., Semple, J., Herman, D., Hong, M.S., Dittmer, S.S., Doddrell, D.M., Toga, A.W., 2003. Dynamics of Gray Matter Loss in Alzheimer’s Disease. The Journal of Neuroscience 23, 994. https://doi.org/10.1523/JNEUROSCI.23-03-00994.2003

Thompson, P.M., Hayashi, K.M., Dutton, R.A., Chiang, M.C., Leow, A.D., Sowell, E.R., de Zubicaray, G., Becker, J.T., Lopez, O.L., Aizenstein, H.J., Toga, A.W., 2007. Tracking Alzheimer’s Disease. Ann N Y Acad Sci 1097, 183. https://doi.org/10.1196/ANNALS.1379.017

Vemuri, P., Gunter, J.L., Senjem, M.L., Whitwell, J.L., Kantarci, K., Knopman, D.S., Boeve, B.F., Petersen, R.C., Jack, C.R., n.d. Alzheimer’s Disease Diagnosis in Individual Subjects using Structural MR Images: Validation Studies.

Vemuri, P., Wiste, H.J., Weigand, S.D., Shaw, L.M., Trojanowski, J.Q., Weiner, M.W., Knopman, D.S., Petersen, R.C., Jack, C.R., 2009a. MRI and CSF biomarkers in normal, MCI, and AD subjects: predicting future clinical change. Neurology 73, 294–301. https://doi.org/10.1212/WNL.0B013E3181AF79FB

Vemuri, P., Wiste, H.J., Weigand, S.D., Shaw, L.M., Trojanowski, J.Q., Weiner, M.W., Knopman, D.S., Petersen, R.C., Jack, C.R., 2009b. MRI and CSF biomarkers in normal, MCI, and AD subjects: Diagnostic discrimination and cognitive correlations. Neurology 73, 287–293. https://doi.org/10.1212/WNL.0B013E3181AF79E5

Vercelletto, M., Martinez, F., Lanier, S., Magne, C., Jaulin, P., Bourin, M., 2002. How to define treatment success using cholinesterase inhibitors. Int J Geriatr Psychiatry 17, 388–390. https://doi.org/10.1002/GPS.612

WHO, 2021. Dementia Fact Sheet [WWW Document]. URL https://www.who.int/news-room/fact-sheets/detail/dementia (accessed 2.15.22).

Yin, C., Li, S., Zhao, W., Feng, J., 2013. Brain imaging of mild cognitive impairment and Alzheimer’s disease. Neural Regen Res 8, 435–444. https://doi.org/10.3969/j.issn.1673-5374.2013.05.007

Young, A.L., Oxtoby, N.P., Daga, P., Cash, D.M., Fox, N.C., Ourselin, S., Schott, J.M., Alexander, D.C., 2014. A data-driven model of biomarker changes in sporadic Alzheimer’s disease. Brain 137, 2564. https://doi.org/10.1093/BRAIN/AWU176

Zanchi, D., Giannakopoulos, P., Borgwardt, S., Rodriguez, C., Haller, S., 2017. Hippocampal and Amygdala Gray Matter Loss in Elderly Controls with Subtle Cognitive Decline. Front Aging Neurosci 9.

